# The impact of antigenic distance on Orthopoxvirus Vaccination and Mpox Infection for cross-protective immunity

**DOI:** 10.1101/2024.01.31.24302065

**Authors:** Jameson Crandell, Valter Silva Monteiro, Lauren Pischel, Zhenhao Fang, Yi Zhong, Lauren Lawres, Luciana Conde, Gustavo Meira de Asis, Gabriela Maciel, Agnieszka Zaleski, Guilherme S. Lira, Luiza M. Higa, Mallery I. Breban, Chantal B. F. Vogels, Nathan D. Grubaugh, Lydia Aoun-Barakat, Alba Grifoni, Alessandro Sette, Terezinha M. Castineiras, Sidi Chen, Inci Yildirim, Andre M. Vale, Saad B. Omer, Carolina Lucas

## Abstract

Immunological memory mediates rapid protection following infection or vaccination including heterologous exposure. However, cross-reactive memory responses in humans remain poorly characterized. We explored the longevity and specificity of cross-protective responses to orthopoxviruses through smallpox vaccination and Mpox virus (MPXV) infection. Smallpox vaccination using Vaccinia virus (VACV)-based vaccines provides a unique opportunity to study long-term cross-protective immunity without antigen re-exposure. We assessed systemic and mucosal responses in four human cohorts, including first-(Dryvax) and/or third-generation (JYNNEOS) smallpox vaccine recipients (vaccinated 1 week-80 years ago), along with Mpox-infected individuals. First-and third-generation smallpox vaccines elicited strong VACV- and MPXV-specific antibodies. VACV-neutralizing antibodies persisted for decades in first-generation vaccine recipients and were further enhanced after JYNNEOS vaccination. However, despite the high levels of anti-MPXV-specific antibodies in the plasma, cross-neutralization activity was directly correlated with the antigenic distance. Higher neutralization was observed for the cowpox virus (CWPXV) than for MPXV, which showed lower antigenic conservation with VACV. Similarly, Mpox-infected patients had lower neutralization titers for VACV than for CWPXV. Individuals who received vaccination boosters showed more robust, diverse, and prolonged cross-neutralizing responses. Long-term memory analysis revealed an increase in neutralization capacity for VACV over decades, with 80-years-old displaying the most robust humoral response, although this trend was not observed for cross-reactive antigens. Finally, T-cell reactivity to VACV and MPXV epitopes was detected decades post-vaccination, suggesting a role of long-lasting cross-reactive T-cell memory responses in vaccine efficacy. Our findings underscore the pivotal influence of antigenic distance on vaccine effectiveness with implications for cross-protective vaccine design.

## Main

The smallpox vaccine is the only vaccine that has led to the eradication of a human disease^1,2^. In this context, VACV has been instrumental as a vaccine and a model for studying antiviral responses. Despite this success, multiple questions regarding its protective mechanisms, cross-protection against other orthopoxviruses, and the durability of these immune memory responses remains unanswered. Specifically, the mobilization of cross-reactive memory cells into a primary immune response, its induction and maintenance over time in comparison to an immunodominant primary response are not fully understood.

Most individuals born before 1980 received smallpox vaccination under the Smallpox Eradication Program (1966-1980)^3^. This program used a variety of live-attenuated strains of VACV, leading to the successful eradication of smallpox and potentially generating broad cross-protective immunity against additional orthopoxviruses, including MPXV^3^. Mpox, previously classified primarily as a zoonotic disease caused by the MPXV virus, has emerged to be the most common orthopoxvirus that infects humans post-smallpox eradication. Although initially endemic to Central and Western Africa, recent outbreaks have demonstrated changes in both transmission patterns and clinical presentations^4,5^. Specifically, mpox was previously described primarily in children in West and Central Africa who had exposure to animals or bush meat with subsequent limited household transmission^6^. In the summer of 2022, there was a mutli-country outbreak of mpox primarily affecting gay, bisexual or other men who have sex with men, and transmission occurring primarily in the sexual setting resulting in genital and rectal lesions at the site of exposure^7,8^

Given the high genetic similarity among poxviruses, it is often assumed that smallpox vaccines can confer broad cross-protection. The first-generation live-attenuated VACV vaccine for smallpox, Dryvax, is believed to provide cross-protective immunity against other orthopoxviruses, including MPXV, but was discontinued due to safety concerns in 2001^9^. Dryvax was replaced by the safer modified vaccinia Ankara (MVA) virus under the brand name JYNNEOS^10^. Although the third-generation smallpox vaccine, JYNNEOS, was reported to have an 86% effectiveness (95% CI 59-95%) for preventing MPXV disease shortly after vaccination and similar estimates after the second dose, detailed insights into the immunogenicity and longevity of its protective responses in humans remain limited^11,12,13^. In this context, we investigated the magnitude and breadth of the cross-protective immunity conferred by smallpox vaccines against MPXV and other poxviruses. Our study leverages the high genetic conservation among orthopoxviruses and the absence of recurrent human exposure to these viruses to explore long-term immunological memory against primary antigens and secondary cross-reactive responses. Decoding these complexities could advance vaccine design, leading to the development of more potent vaccines targeting a broader range of pathogens.

To explore the induction, duration, and cross-reactivity of orthopoxvirus-induced immune memory responses, we assembled four distinct human cohorts. These groups comprised individuals vaccinated with either first- or third-generation smallpox vaccines, along with a third cohort receiving a combination of both vaccines. In addition, we enrolled a fourth cohort of patients with mpox infection. This cohort served as a comparative benchmark for assessing the immune responses elicited by natural infection against those induced by vaccination. To characterize virus-specific immune responses following different smallpox vaccine regimens and natural MPXV infection, we analyzed 335 samples from 219 participants. Cohort 1 (Dryvax Cohort) comprised 78 male and female participants, aged 40-90 years, who were enrolled at the Yale-New Haven Hospital, US, or the Federal University of Rio de Janeiro, Brazil. These individuals received a single dose of the first-generation smallpox vaccine 40-80 years prior to the study (Fig.1a). Plasma and peripheral blood mononuclear cells (PBMCs) samples were collected from November 2022 to June 2023. Cohort 2 (JYNNEOS Cohort) included 62 male and female participants aged > 18 years, who were enrolled at the Yale-New Haven Hospital or the Federal University of Rio de Janeiro. They were administered the third-generation smallpox vaccine, JYNNEOS, in a two-dose regimen with a four-week interval. Plasma and PBMCs were collected longitudinally at baseline (prior to vaccination), 7 and 30-60 days post-first dose, and 7, 30-60, and 180-304 days following the second dose (Fig.1a). Cohort 3 (Dryvax + JYNNEOS), exclusively enrolled at the Federal University of Rio de Janeiro, consisted of 29 male and female participants aged > 40 years old. These individuals received both the first- and third-generation smallpox vaccines. Dryvax was administered at least 40 years prior to this study, whereas the JYNNEOS vaccine was administered only recently. The sampling protocol for Cohort 3 mirrored that for Cohort 2, with longitudinal collections at baseline; 7 and 30-60 days post-first dose; and 7, 30-60 and 180-304 days following the second dose (Fig.1a). Cohort 4, comprising 25 Mpox (MPX)-convalescent patients, was enrolled at the Federal University of Rio de Janeiro from July 2022 to December 2022. Plasma samples were collected 9–80 days after the onset of symptoms (Fig.1a). We examined systemic and mucosal humoral responses (saliva and rectal swabs), neutralizing antibody titers, structural analysis, and T-cell reactivity against VACV, Cowpox virus (CWPXV), and MPXV. The study groups were stratified based on the vaccination regimen, biological sex, and age. Of note, smallpox vaccinations were discontinued in the 1980s: therefore, participants of Cohorts 1 and 3 are aged 40 years or older. In contrast, Cohort 2 primarily consisted of young men, aligning with the target demographics for the JYNNEOS vaccination. To reduce the potential confounding effects of age in our initial analysis, we focused on a subset of younger adults (aged 40-59 years) within Cohorts 1 and 3. Extended-full Cohort 1 was exclusively analyzed in the long-term immunological memory analysis, which involved only participants from Cohort 1 who received Dryvax. Detailed information regarding cohort demographics, vaccination regimens, and infection status is summarized in the Extended Data Table 1, and Extended Data Figure 1.

Plasma antibody reactivity against VACV and MPXV antigens was measured in fully vaccinated individuals from Cohorts 1, 2, and 3. Overall, 85%, 97% to 100% of the analyzed vaccinated participants for Cohorts 1, 2 and 3 respectively, exhibited detectable levels of anti-VACV IgG antibodies in their sera, which target the primary immunodominant antigens present in the vaccine formulations (Fig. 1b). Antibody levels were assessed against key VACV recombinant proteins, B5R, A33R, A27L, and L1R, previously identified as the main targets of neutralizing antibodies^14–16^. These levels were detected across all three vaccine cohorts, with B5R, A33R and A27L antigens eliciting the highest IgG levels. Notably, participants who initially received Dryvax and were recently boosted with JYNNEOS displayed the highest anti-VACV IgG antibody levels (Fig. 1b). No differences were observed in antibody levels between vaccinated participants of different sexes (Extended Data Fig. 2a). Moreover, antibody levels against MPXV-equivalent antigens, B6R, A35R, A29L, E8L, and M1R, were detected in both long-term and recently vaccinated participants compared to non-vaccinated controls, indicating humoral cross-reactivity between VACV and MPXV antigens (Fig. 1b,c). There was no significant difference in the levels of anti-MPXV IgG antibodies between participants who were long-term vaccinated with Dryvax and those recently vaccinated with JYNNEOS regimen, except for the B6R and E8L protein (Fig. 1c). Nevertheless, the most pronounced increase in anti-MPXV IgG antibodies was observed in Dryvax recipients who were recently boosted with JYNNEOS (Fig. 1c).

**Figure 1.**
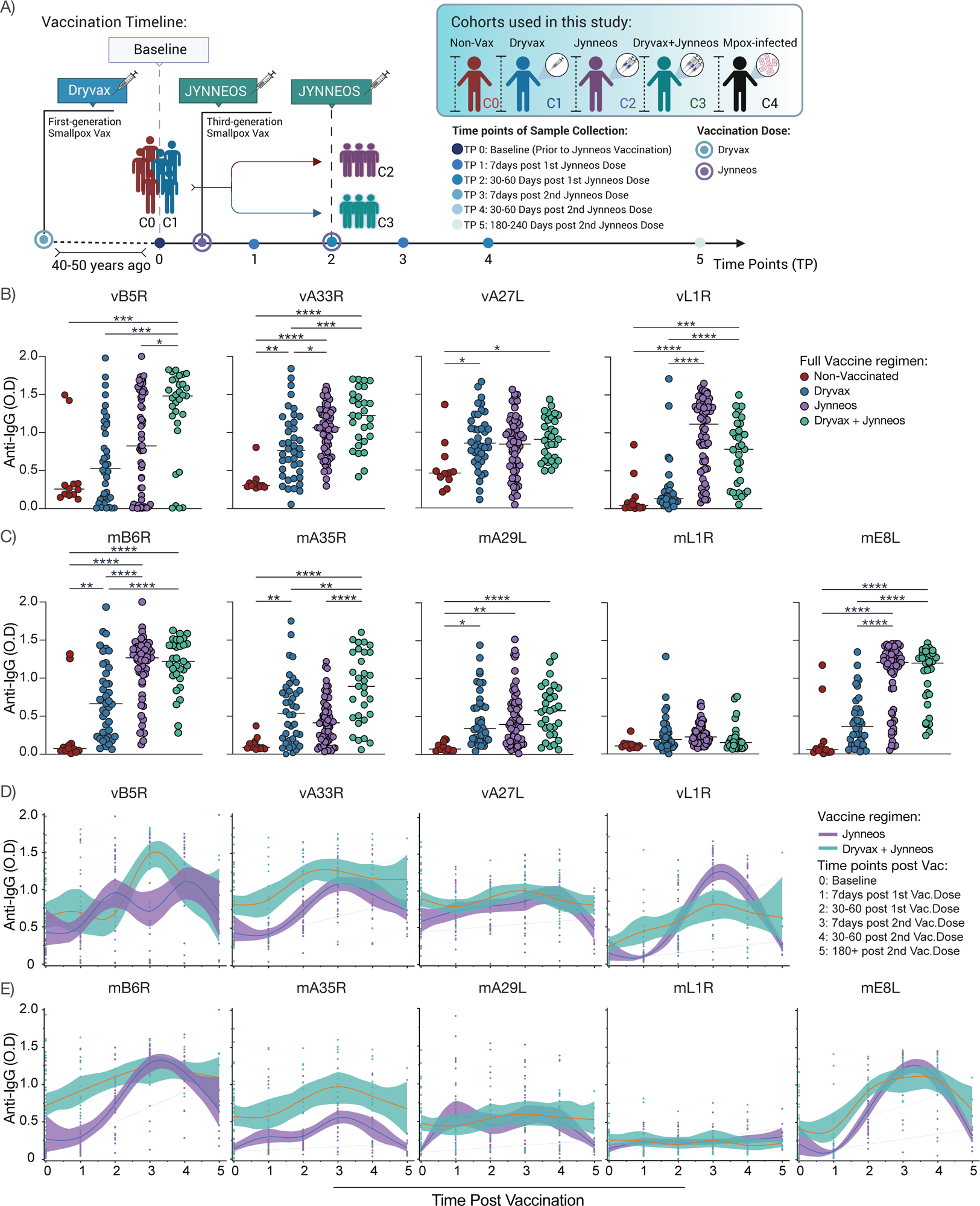
Characterization of Orthopoxvirus-specific antibody responses following first and/or third generation poxvirus vaccines. **a,** Cohort timeline overview indicated by time post vaccination and vaccine regimen received. Participants received Dryvax, JYNNEOS, or both Dryvax and JYNNEOS vaccines and plasma samples were collected at the indicated time points. Participants were then stratified in five main groups: C0, non-vaccinated (C0=24); C1, Dryvax (C1=78); C2, JYNNEOS (C2=62); C3, Dryvax + JYNNEOS (C3=29); C4, Mpox-Infected patients (C4=25). Dryvax was administered as a single dose at least 40 years prior to this study, whereas the JYNNEOS regimen consisted of two vaccination doses and was recently administered to these participants. Created with BioRender. **b,c,** Plasma reactive IgG to viral antigens was measured across all full vaccinated individuals and non-vaccinated controls (C0-C3) (C0=12, C1=40, C2=59, C3=31). **b,** Plasma reactivity to VACV proteins B5R, A33R, A27L, and L1R. **c,** Plasma reactivity to MPXV proteins B6R, A35R, A29L, L1R and E8L. **d,e,** LOWESS regression analysis of virus-specific IgG levels over time following JYNNEOS vaccination. **d,** Plasma reactivity to VACV proteins. **e,** Plasma reactivity to MPXV proteins. Regression lines are shown as purple (JYNNEOS) and green (JYNNEOS + Dryvax); shading represents 95% confidence interval. For the JYNNEOS cohort, baseline controls consisted of non-vaccinated individuals. Baseline controls for the Dryvax + JYNNEOS cohort were individuals who had previously been vaccinated with Dryvax. Significance was assessed by one-way ANOVA corrected for multiple comparisons using Tukey’s method. Baseline: C2=12, C3=40; 7 days post 1^st^ vaccine dose: C2=31, C3=16; 30-60 days post 1^st^ vaccine dose C2=31, C3=14; 7 days post 2^nd^ vaccine dose: C2=30, C3=13; 30-60 days post 2^nd^ vaccine dose: C2=22, C3=14; 180-240 days post 2nd vaccine dose: C2=10, C3=4. Horizontal bars represent median fold change. Non-Vax, non-vaccinated; TP5:180+, 180-240 days. ****P < 0.0001, ***P < 0.001, **P < 0.01, and *P < 0.05.

Next, we investigated the dynamics of antibody responses following vaccination by assessing anti-VACV or anti-MPXV IgG levels longitudinally in the two groups recently vaccinated with the JYNNEOS vaccine: naïve group (JYNNEOS) or those previously vaccinated with Dryvax (JYNNEOS + Dryvax). We found that the third-generation smallpox vaccine, JYNNEOS, induced high levels of virus-specific antibodies in both naïve and previously vaccinated participants. Peak IgG levels against most of the tested viral proteins were observed between 7-30 days after the second vaccination dose (Fig. 1d, e). As expected, virus-specific IgG levels were higher against primary immunodominant antigens (VACV antigens) than against secondary cross-reactive antigens (MPXV orthologs) (Extended Data Fig. 2b, c). Additionally, both anti-VACV and anti-MPXV IgG levels were higher or similar in the group previously vaccinated with Dryvax and subsequently boosted with JYNNEOS, compared to the naïve JYNNEOS-vaccinated group, except for vL1R at the peak of the antibody response (Fig. 1d, e). Antibody levels were found to be dependent on the vaccine dose. Higher virus-specific IgG levels against both VACV and MPXV were observed after the second JYNNEOS vaccination dose in both vaccine-naïve individuals and those previously vaccinated with Dryvax (Extended Data Fig. 3a, b). Notably, virus-specific antibody responses to both primary and cross-reactive secondary antigens quickly declined over time for all viral antigens assessed, except for vL1R, in Dryvax/JYNNEOS-boosted individuals. In contrast to the JYNNEOS naïve group, Dryvax/JYNNEOS boosted individuals had their antibody levels returning to baseline between 6-10 months post-second JYNNEOS vaccination dose (Extended Data Fig. 3c,d). Our data indicate that despite the faster induction of humoral responses to VACV and MPXV viral proteins by the JYNNEOS vaccine in previously vaccinated individuals, these virus-specific antibodies quickly waned after the second vaccination dose, ultimately reaching baseline Dryvax levels within approximately 6-10 months post-vaccination. Overall, these observations indicate that both first- and third-generation vaccines effectively induced antibody responses against VACV and MPXV antigens. This aligns with existing literature, which suggests that first-generation VACV-based vaccines generate long-lasting humoral responses to their primary antigens^9^. Importantly, our data revealed that long-term humoral responses extend to secondary cross-reactive antigens. Our results also revealed that additional booster doses further enhanced these responses.

MPXV is transmitted through respiratory aerosols or via contact with skin lesions^4,5^. Moreover, recent outbreaks have shown genital and rectal lesions in infected patients, which is a unique feature of this virus among all the known orthopoxviruses that infect humans ^4,5^. Due to the virus transmission pattern and this distinctive feature, we asked whether the JYNNEOS vaccine also induced mucosal antibodies. We assessed virus-specific antibody responses in saliva and rectal swabs of individuals recently vaccinated with JYNNEOS. Saliva samples were analyzed between two- and 7-months post-vaccination, whereas rectal swab samples were collected at least six months after the second dose of JYNNEOS. IgA titers were measured against a mixed pool of VACV and MPXV antigens. Virus-specific antibody titers were normalized to the total IgA levels in the respective samples, and the ratio of specificity to total levels was used to estimate the degree of avidity, as indicated in green (Extended Data Fig. 4a). While virus-specific antibodies were detectable in the saliva of JYNNEOS-vaccinated participants, no antibodies were detected in rectal swabs.

Next, we assessed plasma neutralization activity against authentic VACV, CWPX, and MPXV using a 50% plaque-reduction neutralization assay (PRNT50). All vaccination regimens resulted in the production of plasma-neutralizing antibodies targeting the primary immunodominant antigen. Specifically, 71.7%, 97.8%, and 100% of the participants who received one dose of Dryvax, two doses of JYNNEOS, or a combination of three doses (Dryvax followed by JYNNEOS booster), respectively, demonstrated neutralization capacity against VACV (Fig. 2a). Consistent with previous reports^9^, Dryvax induced long-term neutralization responses in individuals vaccinated 40-50 year ago. Participants recently vaccinated with the full JYNEOS regimen exhibited higher neutralization titers against VACV than those in the Dryvax-vaccinated group (Fig. 2a). As predicted by the antibody level analyses (Fig. 1), the most significant increase in neutralization activity was observed in individuals who were previously vaccinated with Dryvax and recently received a JYNNEOS booster, confirming that the neutralization capacity is also dependent on the number of vaccine doses (Fig. 2a and Extended Data Fig. 5a,b). In particular, the neutralizing activity against VACV was directly correlated with anti-L1R, anti-A33R and anti-B5R and antibody titers ((Extended Data Fig. 5c). Overall, the titers of “cross-protective” neutralizing antibodies against related poxviruses, such as CWPX and MPXV, were limited, and directly correlated with the antigenic similarity to VACV (Fig. 2a). Neutralization activity against CWPXV (more antigenically related to VACV) was detected in all vaccinated groups, whereas neutralization against MPXV (more antigenically distant) was only observed in the boosted (Dryvax + JYNNEOS) participants (Fig. 2a-b). Importantly, only individuals with high neutralization titers for VACV (> 2.5) exhibited neutralization titers > 1.5 (exceeding the 1:30 dilution) of MPXV (Extended Data Fig. 5d). In Mpox-infected patients, a similar pattern of antibody cross-reactivity was observed: neutralizing antibody levels correlated with antigenic similarity (MPXV > CWPXV > VACV) (Fig. 2c). However, the overall neutralization titers observed in mpox-infected individuals were lower than those in the vaccinated cohorts. Of note, 52% of individuals in the mpox-infected cohort were immunosuppressed. These observations indicate that antigenic distance and overall antibody levels are determinants of poxvirus-neutralizing cross-reactive antibody responses.

**Figure 2.**
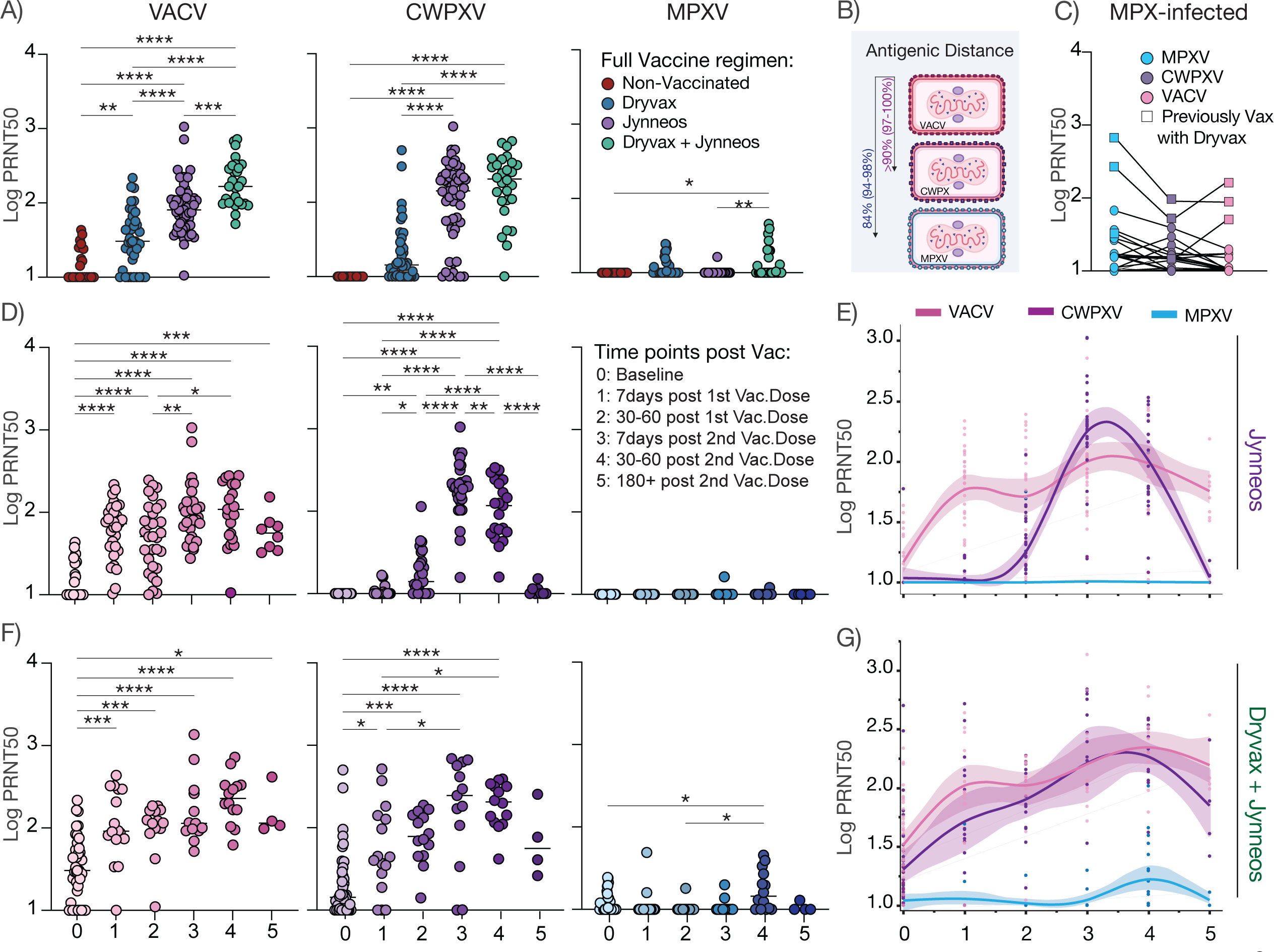
Dynamics of plasma neutralization cross-reactive responses against Orthopoxviruses following vaccination or infection. Blood samples were collected from individuals that received first-generation and/or third generation VACV-based vaccines along with MPXV infected patients. Analysis of immunogenicity were performed using neutralization assays with authentic Vaccinia virus (VACV), Cowpox virus (CWPXV) and Mpox virus (MPXV) virus. **a,** Plasma neutralization titers against VACV, CWPXV and MPXV across full vaccinated individuals and non-vaccinated controls (C0=21, C1=40, C2=55, C3=28). **b,** Schematic figure illustrating antigenic similarity from VACV across Orthopoxviruses used within this study. Colors indicate different viruses, VACV (pink), CWPXV (purple), MPXV (blue) and arrows indicate antigenic distance. Full genomic similarity is indicated outside the parentheses, while the percentage of genomic similarity across the eight immunogenic proteins, which are the main targets for neutralization, is specified within the parentheses. Created with BioRender. **c,** Plasma neutralization capacity against VACV, CWPXV, and MPXV in MPXV-infected patients (C4=25). Each dot represents a single patient. Squares represent patients that were previously vaccinated with Dryvax vaccine. **d,e,f,g,** Plasma neutralization capacity against VACV, CWPXV and MPXV over time following JYNNEOS vaccination. **d,** Neutralization titers over time in naïve, non-vaccinated individuals after the JYNNEOS regimen. **e,** LOWESS regression comparisons of plasma neutralization capacity against each orthopoxvirus over time in naïve, individuals, following the JYNNEOS regimen. **f,** Neutralization titers over time after the JYNNEOS regimen, in individuals previously vaccinated with Dryvax. **e**, LOWESS regression comparisons of plasma neutralization capacity against each orthopoxvirus over time following the JYNNEOS regimen, in individuals previously vaccinated with Dryvax. Regression lines are shown as pink (VACV), purple (CWPXV) and light blue (MPXV); shading represents 95% confidence interval. Significance was assessed using mixed-effect analysis corrected for multiple comparisons using by one-way ANOVA corrected for multiple comparisons using Tukey’s method. Baseline: C2=22, C3=40; 7 days post 1^st^ vaccine dose: C2=31, C3=16; 30-60 days post 1^st^ vaccine dose C2=31 C3=14; 7 days post 2^nd^ vaccine dose: C2=30, C3=13; 30-60 days post 2^nd^ vaccine dose: C2=21, C3=14; 180-240 days post 2nd vaccine dose: C2=8, C3=4. Lines indicate neutralization dynamics across vaccine regimen. Horizontal bars represent average ± SD. TP5:180+, 180-240 days. ****P < 0.0001, ***P < 0.001, **P < 0.01, and *P < 0.05.

To assess the magnitude and duration of both primary and cross-reactive secondary neutralization responses following JYNNEOS vaccination, we longitudinally measured the neutralization capacity of orthopoxviruses in naïve, non-vaccinated, and Dryvax-vaccinated individuals. Consistent with the systemic antibody level analysis, the peak of neutralization titers for both VACV and CWPXV were observed after the second JYNNEOS vaccination dose in both groups (Fig. 2d,e). Notably, neutralizing titers against MPXV were only detected 30-60 days post-second JYNNEOS dose in those previously vaccinated with Dryvax (Fig.2f,g). The number of vaccine doses affected not only the magnitude and breadth of the response—with individuals receiving three doses exhibiting the highest titers against the primary immunodominant antigen and cross-reactive CWPX and MPXV—but also the kinetics of antibody responses against secondary cross-reactive viruses (Fig.2d-g). Specifically, in naïve, non-vaccinated individuals, a single JYNNEOS dose led to a rapid and sustained increase in neutralization titers against the primary antigen, VACV. Two JYNNEOS doses in these individuals induced increased neutralization titers against both VACV and the closely related CWPXV. Importantly, neutralization titers against CWPXV quickly declined to baseline levels, 6-10 months post vaccination dose 2 (Fig.2d and Extended Data Fig. 5e, f). No neutralization titers against MPXV were observed in these individuals (Fig.2d and Extended Data Fig. 5e). Notably, individuals previously vaccinated with Dryvax and subsequently receiving their first JYNNEOS dose (totaling two doses) exhibited similar antibody dynamics for VACV and CWPX (Fig.2f). Additionally, these individuals (1 dose Dryvax+ 1 dose JYNNEOS) had neutralizing antibody dynamics similar to naïve individuals who received two JYNNEOS doses, demonstrating a rapid and sustained neutralization capacity against VACV and CWPX (Fig.2f,g and Extended Data Fig. 5e,f). Moreover, after the third dose (second JYNNEOS dose), these boosted individuals showed a significantly higher neutralization capacity against MPXV (Fig.2f,g). Neutralization titers against MPXV quickly waned over time, returning to baseline levels within 6-10 months after vaccination (Fig.2f,g and Extended Data Fig. 5f). Importantly, we next examined whether varying the time interval between JYNNEOS vaccine doses would affect the antibody neutralization capacity. JYNNEOS was implemented as a two-dose vaccine with a standard 4-week interval between doses. No significant differences in neutralizing antibody titers against VACV or MPXV were observed when the interval was extended (>41 days). Interestingly, an increase in neutralization titers for CWPXV was noted in previously non-vaccinated individuals who received the second JYNNEOS dose after a longer interval (Extended Data Fig. 5g). Our data indicate that both first- and third-generation smallpox vaccines induce potent neutralizing responses against VACV. However, despite the detection of high levels of anti-MPXV-specific antibodies in the plasma, neutralization activity directly correlates with viral antigenic distance, with a poor neutralizing response observed for MPVX, in contrast to CWPXV. Additionally, the number of vaccine boosters affects not only the magnitude but also the breadth and kinetics of the response, with individuals who received three vaccination doses displaying the most robust, diverse, and prolonged neutralization responses against cross-reactive viruses.

To better understand the observed variations in antibody neutralization titers, we investigated the genetic differences between VACV immunogenic proteins and their orthologs in CWPXV and MPXV 2022 isolates. Sequence analysis was conducted for five VACV-immunogenic proteins: B6R, A35L, A29L, M1R, and E8L. These proteins elicit strong neutralizing antibody responses and are primary neutralization targets of VACV. The VACV reference antigen sequence exhibited high genetic similarity, ranging from ∼94% to ∼100%, with Mpox and Cowpox virus antigens, confirming that these immunogenic proteins are highly conserved among orthopoxviruses. Importantly, despite the overall high genetic identity, CWPXV antigens have higher sequence similarity with VACV proteins than MPXV orthologs (Extended Data Fig. 6a,b). We identified all residues (highlighted in red) that are identical for CWPXV and VACV but different for MPXV (Extended Data Fig. 6a). The highest genetic similarity was observed for L1R antigens (∼99-100%), with A27 (∼94%-98%) demonstrating the lowest sequence similarity (Extended Data Fig. 6b). Next, we performed structural analysis to evaluate whether the mutations observed exclusively for MPXV were located within the neutralizing epitope regions, potentially accounting for the observed variation in neutralization activity (Extended Data Fig. 6c-f). Most homolog mismatches identified in the sequence alignment do not overlap with neutralization epitope regions. However, certain residues on the mpox sequence were adjacent to the neutralization epitope region, including Q117K and L118S in A35R and T146M, L66I, and S65T in E8L (Extended Data Fig. 6c-f). Therefore, these residues may affect the binding of neutralizing antibodies. The above observations indicate that the potent virus-specific humoral responses elicited by first- and third-generation VACV-based vaccines act through conserved antigen epitopes. However, the homolog mismatches also suggest low cross-protective neutralizing antibody responses against distantly related poxviruses, particularly MPXV. Despite the high sequence similarity among VACV, CWPXV, and MPXV orthologs, we found specific amino acid mismatches present exclusively in mpox antigens located near neutralization sites that may contribute to antibody evasion.

Immunological memory can be maintained in an antigen-dependent or -independent manner, with the latter not requiring repeated antigenic boosting event^17,18^. The intricacies of immunological memory maintenance, including the mobilization of cross-reactive memory cells into a primary immune response and its subsequent sustenance compared to an immunodominant primary response, remain unclear. We next assessed the persistence of immune memory in individuals vaccinated long ago with the Dryvax vaccine. To address this, we expanded our initial cohort to include participants aged 60-89 years old (Fig.3a). Given that all participants from Cohort 1 reported receiving their smallpox vaccines during childhood (ages 0-10 years), and considering the self-reported nature of vaccination dates, we chose to stratify participants by age decades rather than by time since vaccination. The persistence of humoral responses over time was evaluated using ELISA and neutralization assays. Strikingly, anti-VACV and anti-MPXV IgG levels were detectable several decades after smallpox vaccination (Fig.3b). Particularly, robust antibody responses against A33/A35 and E8L were observed. A trend was also observed for anti-IgG levels against the B6/B5R antigens. Additionally, the levels of anti-A27/A29 and anti-L1R were stable over the decades (Fig.3b). Our data also revealed that the cross-reactive humoral response to MPXV orthologs was similar or slightly lower than that to VACV antigens (Fig.3b). The kinetics of antibody production against homologous and heterologous antigens over time were similar and long lasting (Fig.3b). We also evaluated the neutralizing activity against VACV, CWPXV, and MPXV over time. Neutralizing titers against VACV also displayed a gradual increase over the decades, suggesting an improvement in antibody affinity over time. Conversely, neutralizing titers against CWPXV and MPXV did not exhibit the same dynamics and maintained stable levels (Fig. 3c). These findings indicate that the increase in neutralization titers is specific to the primary immunodominant response and does not extend to cross-reactive secondary antigens. Our data also point to an unexpected improvement in the antibody neutralization capacity to overcome immune senescence, with individuals aged 80 years showing the most robust humoral response against VACV.

**Figure 3.**
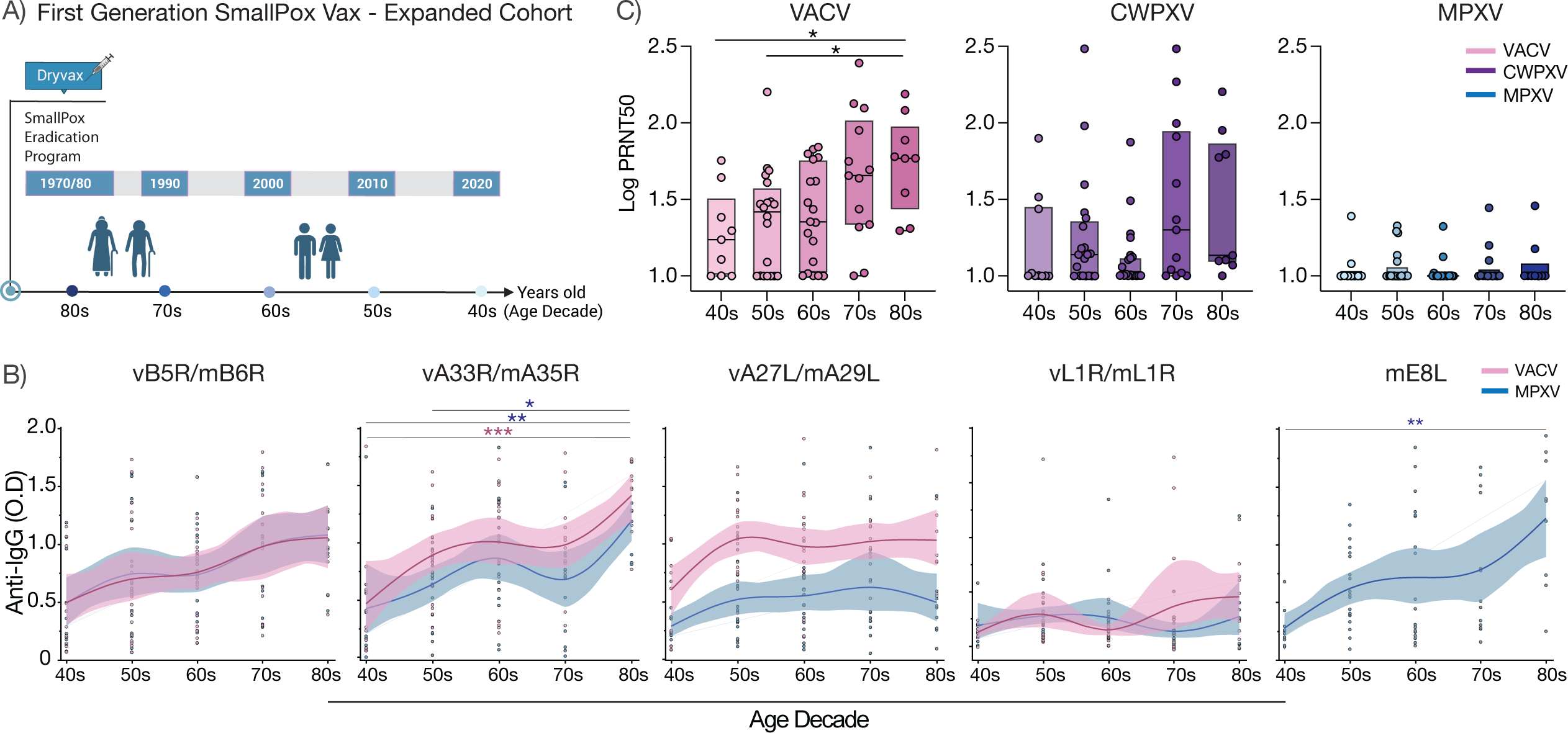
Long-term cross-reactive humoral memory response following Dryvax vaccine. **a,** Schematic figure of expanded Dryvax cohort. Participants vaccinated before the US Smallpox Eradication campaign ended (∼1970) were recruited on the study and categorized by their birth decade. Samples were collected from participated vaccinated 40-80years prior to this study. **b,** Plasma reactivity to VACV and MPXV proteins (B5/6R, A33/35R, A27/29L, L1/M1R, and E8L) in Dryvax vaccinated individuals over decades. Regression lines indicate plasma IgG levels anti-VACV antigens (pink) or anti-MPXV antigens (blue); shading represents 95% confidence interval. **c,** Plasma neutralization titers against VACV, CWPXV, and MPXV over decades. Significance was assessed by one-way ANOVA corrected for multiple comparisons using Tukey’s method. 40s=9; 50s=20; 60s=19; 70s=13; 80s=9. Boxes represent average ± SD.****P < 0.0001, ***P < 0.001, **P < 0.01, and *P < 0.05.

Previous research on VACV has demonstrated that antibody responses are crucial for disease prevention, whereas T cell responses are important for controlling and terminating poxvirus infections^19,20^. Despite more than 94-98% genomic identity between VACV and MPXV immunogenic proteins, our findings suggest that both the first- and third-generation smallpox vaccines do not induce potent levels of neutralizing antibodies against MPXV, raising the possibility that T cells play a role in the cross-protective responses. Hence, we analyzed cross-reactive T cell responses against MPXV and other orthopoxviruses using flow cytometry in our established cohorts. T cell responses were evaluated for their ability to recognize orthopoxvirus peptide pools (OPX) or a specific Mpox peptide pool (MPX). The responses were assessed using activation-induced marker (AIM) assays, as previously described^21^. To detect low-frequency peptide-specific T cell populations, we first expanded antigen-specific T cells by stimulating PBMCs from vaccinated individuals with OPX or MPX peptide pools ex vivo for nine days, followed by restimulation with the same peptide pools. Consistent with previous data in literature^22^, we observed that JYNNEOS vaccination led to an increase in OPX-reactive CD4 and CD8 T cells, as evidenced by the upregulation of activation markers, including OX-40, CD69, and CD137 (Fig.4a). Furthermore, we found that both first- and third-generation VACV-based vaccines induce T cell responses that cross-recognize MPXV-derived epitopes (Fig.4a). Such cross-reactivity was confirmed in both recently JYNNEOS and long-ago Dryvax-vaccinated individuals, including those who received Dryvax vaccines over 80 years ago (Fig.4b). Thus, our data indicate that smallpox vaccines effectively induce long-term T cell responses to primary VACV antigens, as well as long-term cross-reactive responses to MPXV.

**Figure 4.**
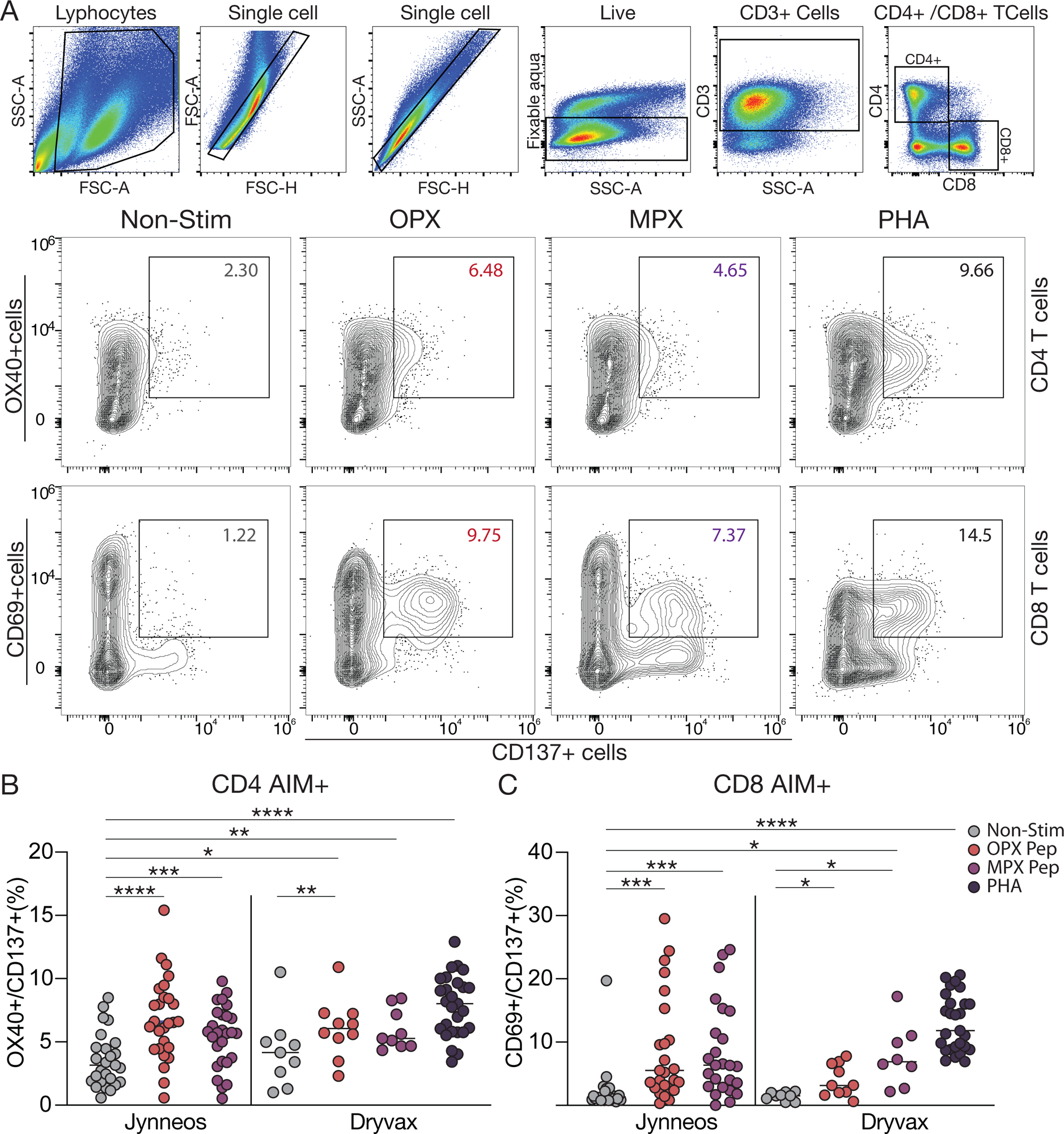
Cross-reactive T cells responses following first and/or third generation poxvirus vaccines. Orthopoxviruses-reactive CD4+ and CD8+ T cells after in vitro PBMC stimulation with orthopoxviruses peptide pools (OPX) or Mpox specific peptide pool (MPX)**. a,** Gating strategy and representative dot plots showing the percentage of double-positive cells expressing OX40+CD138+ (AIM+) CD4+ T cells (top) and CD69+CD138+ (AIM+) CD8+ T cells (bottom). **b,** Analysis of AIM+ CD4+ T cells and AIM+ CD8+ T cells in individuals recently fully vaccinated with JYNNEOS compared to those vaccinated with Dryvax long ago. Significance was assessed by one-way ANOVA corrected for multiple comparisons using Tukey’s method. JYNNEOS: CD4=26; CD8=24. Dryvax: CD4=10; CD8=10. Each dot represents a single individual. Horizontal bars represent average ± SD. Stim, non-stimulated cells; PHA, phytohemagglutinin was used as positive control. ****P < 0.0001, ***P < 0.001, **P < 0.01, and *P < 0.05.

## Discussion

In the wake of the 2022-2023 Mpox outbreak, investigations into smallpox vaccine effectiveness have been essential for evaluating pandemic preparedness and our ability to curb future orthopoxvirus outbreaks. Due to the highly conserved nature of orthopoxviruses, smallpox vaccines are believed to be cross-protective against other orthopoxviruses, such as Mpox, leading to the authorization of both ACAM2000 and JYNNEOS to be used prophylactically against Mpox infection. However, without thorough evaluation of cross-protection and memory responses elicited from third-generation smallpox vaccines, there are substantial gaps in our understanding of the effectiveness and duration these vaccines may have in protecting against related orthopoxvirus infections. This study addressed these gaps by investigating the cross-reactive immune responses to orthopoxviruses across cohorts of patients vaccinated with Dryvax, or/and JYNNEOS, or Mpox infected. We observed that these vaccines elicit long-lasting humoral and cell-mediated memory responses that cross-protect as an inverse function of antigenic distance such that as antigenic distance increased cross-protection declined. Additionally, we found that booster vaccination doses are associated with increased breadth and sustainment of cross-reactive immunological memory responses.

Long-lasting cross-reactive immunity to MPXV has previously been described in mouse models and was recently described in Dryvax-vaccinated individuals exhibiting both VACV-reactive and MPXV-neutralizing antibodies^23,24^. Zaeck et al. also observed a significant boost in antibody responses and MPXV neutralization upon administration of three doses of VACV-based homologous vaccines^23^. By studying key orthopoxvirus antigens, our study confirms and expands these findings by indicating that neuralization capacity was associated with both antibody titer and antigenic distance from the primary target antigen in VACV. Importantly, by evaluating the kinetics of the vaccine response, our study helps deconvolute the role of VACV-based vaccine boosts in generating antibody responses targeting primary (VACV) *versus* cross-reactive (CWPXV and MPXV) antigens. Whereas JYNNEOS boost in Dryvax-vaccinated individuals only generated a transient (<10 months) humoral response to primary VACV antigens, additional boosts were associated with long lasting improvement of cross-reactive humoral responses^25^.

The observations made in this study that different VACV-based vaccination strategies result in minimal neutralization capacity to the 2022 USA Mpox strain, along with findings from Zaeck et al., contradicts earlier reports anticipating high cross-protection against Mpox from VACV-based smallpox vaccines^26,27^. Our structural and antigen-binding based analyses suggest that reduced Mpox responses are likely attributed to lower homology between VACV and the Mpox strain (84% total genome and 94-98% for the immunogenic proteins)^28^. In fact, multiple analyses, including plasma from Mpox-infected individuals, point to an inverse relationship between cross-protection and antigenic similarity^29,30^. This “antigenic distance” phenomenon has been reported in other viruses, including influenza, in which studies suggest a negative interference in mounting novel immune responses when antigenic distance is small between seasonal vaccines^31^. Mechanistically, it was proposed that homologous boosting in SARS-CoV-2 could expand antibody breadth by enhancing subdominant antigen availability through a process of immunodominant epitope masking^32^. This was also addressed recently by Schiepers et al. through molecular fate mapping of secreted antibodies, which revealed that antibody responses to sequential homologous boosting mostly derive from primary B cells, an effect that drastically decreases as a function of antigenic distance^33^. Conversely, when antigenic distance is large, cross-reactive immune responses are diminished posing significant challenges for developing effective vaccines able to overcome “the original antigenic sin”^34^. Therefore, it is possible that the observed improvement in Mpox cross-reactive humoral responses following the third dose is merely a result of increased titers for antibodies with a small capacity of cross-protection. Alternatively, sequential boosts could have triggered increased breadth due to epitope masking. Regardless, our structural and antigen-binding-based analyses could be informative in understanding this phenomenon of antigenic distance within poxviruses and in the development of more effective vaccines.

Analysis of long-lasting humoral responses was achieved through extension of our Dryvax cohort to include individuals ≥ 40 years that were previously vaccinated with Dryvax. Similar to previous reports, we observed detectable antibody responses in patients vaccinated up to 80+ years ago^9,35^. Strikingly, by stratifying patients according to age decade, we also observed that neutralization capacity against both primary (VACV) and secondary (MPXV) vaccine targets significantly increased with age. One recent study has described the capacity for long-primed germinal centers lasting for at least 6 months in rhesus macaques which was ascribed to prolonged affinity maturation and clonal migration of memory B cells in response to the HIV Env protein^36^. Similarly, another recent study described increased somatic hypermutation and potency of antibodies up to 6.2 months post infection with SARS-CoV-2 in response to antigen persistence^37^. However, until now, gradual affinity maturation across multiple decades has yet to be reported and could suggest that antibody response to first-generation smallpox vaccination continues to improve over multiple decades - potentially overcoming senescence, though further investigation into this mechanism is needed.

Investigations into cell-mediated memory responses were conducted in tandem in patients vaccinated with Dryvax or/and JYNNEOS. Using similar methods to those previously described, patient T cells were stimulated with either OPXV- or MPXV-specific peptide pools^21,22^. Through this, we observed memory T cell responses to both VACV and MPXV up to 60 years post-vaccination with Dryvax, indicating that lasting, cross-reactive, cell-mediated responses are elicited from first-generation smallpox vaccines ^31^. Additionally, we expand upon previous reports by demonstrating that both Dryvax and JYNNEOS evoke cross-reactive cell-mediated responses to both VACV and MPXV. While previous studies have suggested potential for cross reactive T cell responses, we provide a more comprehensive understanding of induced cell-mediated responses from both first- and third-generation smallpox vaccines to broad orthopoxvirus and Mpox-specific peptide pools and the longevity of this response^21,37^.

Limitations in this study include heterogeneity within the cohorts. Due to the distinctive nature of the administration of the Dryvax and JYNNEOS vaccines, populations within these two cohorts vary widely. Additionally, in the mucosal antibody analysis only saliva samples were able to be collected within biologically relevant timepoints (1-6months post vaccination) with rectal swab samples only becoming available much later resulting in samples 6+ months post vaccination. Furthermore, as previously described, the use of the peptide pools may not reflect physiologically relevant targets; to date the MPXV pools have yet to be validated, hindering our ability to definitively confirm cross-protective cell-mediated responses^21^. Finally, the AIM+ assay for analyzing T cell response was used on cells post 9 day in vitro stimulation which may have affected the expression of membrane markers.

## Data Availability

All data produced in the present study are available upon reasonable request to the authors.

## Methods

### Ethic Statement

This study was approved by the Yale Human Research Protection Program Institutional Review Board (IRB protocol ID 2000033415) and the Research Ethics Committee of the HUCFF/UFRJ (protocol number CAAE 62281722.5.0000.5257). Our study encompasses four human cohorts, comprising individuals vaccinated with first and/or third-generation smallpox vaccines, as well as patients infected with Mpox. Informed consent was obtained from all enrolled vaccinated/infected individuals. None of the participants experienced serious adverse effects after vaccination.

### Study Participants

For this study we enrolled 218 participants from the United States and Brazil, yielding 335 samples, including PBMCs, plasma, saliva, and rectal swabs. The participants were categorized into four cohorts based on their vaccination/infection status (Fig.1a): Cohort 1 comprises 78 individuals vaccinated with the first-generation smallpox vaccine, Dryvax, 40-80 years ago; individuals reported no prior infection or direct contact with infected patients. Samples were collected at a single time point. Cohort 2 consists of 62 individuals recently vaccinated with the third-generation smallpox vaccine, JYNNEOS. Longitudinal samples were collected at baseline (pre-vaccination), 7 and 30-60 days after the first dose, and 7, 30-60, and 180-304 days post the second dose. Cohort 3 included 29 individuals vaccinated with both Dryvax (40-80 years ago) and recently with JYNNEOS, with sampling mirroring Cohort 2. Cohort 4 comprises 25 MPXV-infected patients, with samples collected 9–80 days post symptom onset. Demographic data were gathered via a screening questionnaire at blood collection and electronic health records, contributing to (Extended Data Table 1 and Extended data Figure 1). Clinical data collection used EPIC EHR (May 2020) and REDCap 9.3.6. A separate team managed blood acquisition, and ELISA, neutralization, and flow cytometry analyses were performed in a blinded manner.

### Isolation of Plasma and PBMCs

Whole blood was collected in heparinized CPT blood vacutainers (BDAM362780, BD) and kept on gentle agitation until processing. All blood was processed on the day of collection in a single step standardized method. Plasma samples were collected after centrifugation of whole blood at 600*g* for 20□min at room temperature without a break. The undiluted plasma was transferred to 1.8-ml cryogenic vials (V7884-450EA, Sigma Aldrich), aliquoted, and stored at −80□°C for subsequent analysis. The PBMC layer was isolated according to the manufacturer’s instructions. Cells were washed twice with PBS before counting. Pelleted cells were briefly treated with ACK lysis buffer for 2□min and then counted. Percentage viability was estimated using standard trypan blue staining and an automated cell counter (AMQAX1000, Thermo Fisher). PBMCs were stored at −80□°C for subsequent analysis.

### Orthopoxvirus Specific Antibody Measurements

MaxiSorp plates (96 wells; 442404, Thermo Scientific) were coated with 50□μl per well of either recombinant MPox A35R (A3R-M52H3-100 μg, ACROBiosystems), MPox A29L (A2L-M52H3-100 μg, ACROBiosystems), MPox E8L (E8L-M52H3-50 μg, ACROBiosystems), MPox L1R (L1R-M5241-50 μg, ACROBiosystems), MPox H3L (H3L-M52H1-50 μg, ACROBiosystems), MPox A30L (A3L-M5243-50 μg, ACROBiosystems), VACV A33R (40896-V07E-100 μg, Sino Biological), VACV A27L (40897-V07E-100 μg, Sino Biological), VACV L1R (40903-V07H-100 μg, Sino Biological), or VACV OPC005 [vD8L] (CSB-EP322653VAA-100 μg, CusaBio) at a concentration of 2□μg/ml in PBS and were incubated overnight at 4□°C. The coating buffer was removed, and plates were incubated for 1□h at room temperature with 200□μl of blocking solution (PBS with 0.1% Tween-20 and 3% milk powder). Plasma was diluted at 1:100 in dilution solution (PBS with 0.1% Tween-20 and 1% milk powder), and 100□μl of diluted serum was added for 2 h at room temperature. Plates were washed three times with PBS-T (PBS with 0.1% Tween-20) and 50□μl of HRP anti-human IgG antibody (1:5,000; A00166, GenScript) diluted in dilution solution added to each well. After 1□h of incubation at room temperature, plates were washed six times with PBS-T. Plates were developed with 100□μl of TMB Substrate Reagent Set (555214, BD Biosciences) and the reaction was stopped after 5□min by the addition of 2 N sulfuric acid. Plates were then read at a wavelength of 450□nm.

### Mucosal and Total IgA Antibody Measurement

To measure mucosal antibody response, virus-specific antibodies were calculated in proportion to total IgA from each sample. In this way, two ELISAs were conducted in tandem (total IgA and poxvirus specific), the methods used are as follows. MaxiSorp plates (96 wells; 442404, Thermo Scientific) were coated with 50□μl per well of either unlabeled anti-IgA (Goat Anti-Human IgA, Mouse/Bovine/Horse SP ads-UNLB, 2053-01, SouthernBiotech) at 1 µg/mL or a mixture of poxvirus specific proteins (MPox A35R (A3R-M52H3-100 μg, ACROBiosystems), MPox A29L (A2L-M52H3-100 μg, ACROBiosystems), MPox E8L (E8L-M52H3-50 μg, ACROBiosystems), VACV A33R (40896-V07E-100 μg, Sino Biological), VACV A27L (40897-V07E-100 μg, Sino Biological), and VACV L1R (40903-V07H-100 μg, Sino Biological)) at a concentration of 1 µg/mL per protein; plates were incubated overnight at 4 °C. The coating buffer was removed, and plates were incubated for 1□h at room temperature with 200□μl of PBS with 1% Bovine Serum Albumin (BSA). During incubation, samples were thawed and used at room temperature; all samples were centrifuged at 14000rpm prior to dilution in PBS-1% BSA. For saliva total IgA measurements, samples were diluted serially 1:40 to 1:87480; similarly, for poxvirus specific, samples were diluted serially 1:10 to 1:21870. For rectal swab samples, both total IgA and poxvirus-specific measurements were diluted serially from 1:1 to 1:2187. Standards (Human IgA Kappa-UNLB, 0155K-01, SouthernBiotech) were serially diluted 1:100 to 1:72900; standard curves started at 1 µg/mL for rectal swab samples and 0.1 µg/mL for saliva samples. 50 µL of diluted sample/standard was added to each well and incubated for 2h at room temperature. After incubation, plates were washed five times with 200 µL of PBS per well and 50□μl of HRP anti-human IgA antibody (Goat Anti-Human IgA, Mouse/Bovine/Horse SP ads-HRP, 2053-05, SouthernBiotech) diluted 1:5000 in PBS-1%BSA was added to each well. After 1□h of incubation at room temperature, plates were washed five times with 200 µL PBS-1%BSA per well. Plates were developed with 50□μl per well of TMB Substrate Reagent Set (555214, BD Biosciences) and the reaction was stopped according to development of the standard curve (approx. 4-5 min) by the addition of 2 N sulfuric acid. Plates were then read at a wavelength of 450□nm.

### T Cell Stimulation

For the in vitro stimulation, PBMCs were stimulated with Orthopox and MPox-specific peptide pools at the concentration of 1□μg ml^−1^ per peptide and cultured for 9 days. On day 0, PBMCs were thawed, counted, and plated in a total of 5–8□×□10^5^ cells per well in 200□ul of RPMI 1640 medium (Gibco) supplemented with 1% sodium pyruvate (NEAA), 100□U/ml penicillin–streptomycin (Biochrom) and 20% FBS at 37□°C and 5%□CO_2_. On day 1, cells were washed and stimulated with peptide megapools (MP) identified as: OPXV CD4, OPXV CD8, MPXV CD4, and MPXV CD8 as previously described^21^. Stimulation controls were performed with RPMI 1640 medium (Gibco) supplemented with 1% sodium pyruvate (NEAA), 100□U/ml penicillin–streptomycin (Biochrom) and 20% FBS as a negative control and Phytohemagglutinin (PHA) in complete RPMI1640 as a positive control. Peptide pools were used at 1□μg ml^−1^ per peptide. Incubation was performed at 37□°C, 5%□CO_2_ for 9 days. On days 3, 5, and 7, cells were fed with a cocktail of cytokines to support cell survival. The cytokines used includes: 10 IU ml^−1^ IL-2 (Miltenyi Biotec, 130097743), 10 ng ml^−1^ IL-7 (R&D Systems, 207-IL-025), 10 ng ml^−1^ IL-15 (PeproTech, 200-15), 1000 IU ml^−1^ GM-CSF (PeproTech, 300-03), 500 IU ml^−1^ IL-4 (R&D Systems, 204-IL-100), and 100 ng ml^−1^ Flt3-L (R&D Systems, 308-FKN-025/CF). On day 9, cells were restimulated with 1□μg ml^−1^ per peptide and subsequently incubated for 12□h, with the last 4□h being in the presence of 10□μg ml^−1^ brefeldin A (Sigma-Aldrich) for intracellular staining. Following this incubation, cells were washed with PBS 2□mM EDTA and prepared for analysis by flow cytometry.

### Flow Cytometry

Antibody clones and vendors were as follows: BV605 anti-hCD3 (UCHT1, 1:300; BioLegend), BV785 anti-hCD4 (SK3, 1:300; BioLegend), BV421 anti-hCCR7 (G043H7, 1:50; BioLegend), AF700 anti-hCD45RA (HI100, 1:100; BioLegend), APC anti-hCD69 (FN50, X; BioLegend), BV711 anti-hCD137 (4B4-1, X; BioLegend), FITC anti-hGranzyme B (GB11, X; BioLegend), APC/Fire750 anti-hCD8 (SK1, X; BioLegend), PE anti-hCD38 (S17015A, X; BioLegend), PE anti-hIFNγ (4S.B3, X; BioLegend), BB515 anti-hHLA-DR (G46-6, X; BD Biosciences), BB700 anti-hCD134 (ACT35, X; BD Biosciences), PE-Cy7 anti-hCD127 (HIL-7R-M21, X; BD Biosciences), BV785 anti-hCD19 (SJ25C1, 1:300; BioLegend), BV421 anti-hCD138 (MI15, 1:300; BioLegend), AlexaFluor700 anti-hCD20 (2H7, 1:200; BioLegend), AlexaFluor 647 anti-hCD27 (M-T271, 1:350; BioLegend), PE/Dazzle594 anti-hIgD (IA6-2, 1:400; BioLegend), BV711 anti-hCD38 (HIT2, 1:200; BioLegend), AlexaFluor 700 anti-hTNFa (MAb11, 1:100; BioLegend). In brief, freshly isolated PBMCs were plated at 1–2□×□10^6^ cells per well in a 96-well U-bottom plate. Cells were resuspended in Live/Dead Fixable Aqua (Thermo Fisher) for 20□min at 4□°C. Following a wash, cells were blocked with Human TruStan FcX (BioLegend) for 10□min at room temperature. Cocktails of desired staining antibodies were added directly to this mixture for 30□min at room temperature. For secondary stains, cells were first washed and supernatant aspirated; then to each cell pellet, a cocktail of secondary markers was added for 30□min at 4□°C. Before analysis, cells were washed and resuspended in 100□μl 4% PFA for 30□min at 4□°C. Following this incubation, cells were washed and prepared for analysis on an Attune NXT (Thermo Fisher). Data were analysed using FlowJo software version 10.6 software (Tree Star).

### Cell Lines and Viruses

HeLa cells (ATCC® CCL-2.2) were cultured in DMEM supplemented with 1% sodium pyruvate and 10% FBS at 37□°C and 5%□CO2, similarly BS-C-1 cells (ATCC® CCL-26) were cultured in MEM supplemented with 1% sodium pyruvate and 10% FBS at 37 °C and 5% CO2. The cell line was obtained from the ATCC® and tested negative for contamination with mycoplasma. Vaccinia Virus, Modified Vaccinia Ankara (MVA) was obtained from BEI Resources (no. NR-1) and was amplified on HeLa cells. Similarly, Monkeypox Virus, hMPXV/USA/MA001/2022 (Lineage B.1, Clade IIb) and Cowpox Virus, Brighton Red were obtained from BEI Resources (no. NR-52281 and NR-88, respectively) and amplified in BS-C-1 cells. The re-sequenced genomes were submitted to the National Center for Biotechnology Information (NCBI; GenBank accession numbers: U94848, ON563414, and NC_003663, respectively). Cells were infected at a multiplicity of infection of 0.01 for 4 days to generate a working stock, and after incubation, the supernatant was clarified by centrifugation (450g for 5□min) and filtered through a 0.70-µm filter. The pelleted virus was then resuspended in media respective to cell type and aliquoted for storage at −80□°C. Viral titers were measured by standard plaque assay using HeLa for VACV and BS-C-1 for CPXV and MPXV. In brief, 300□µl of serial fold virus dilutions were used to infect HeLa or BS-C-1 cells in MEM supplemented with NaHCO3, 4%□FBS and 0.6% Avicel RC-581. Plaques were resolved at 90□h post-infection by fixing in 10% formaldehyde for 1□h followed by 0.5% crystal violet in 20% ethanol staining. Plates were rinsed in water to plaques enumeration. All MPXV experiments were performed in a biosafety level 3 laboratory with approval from the Yale Environmental Health and Safety office.

### Neutralization Assays

Sera from vaccinated volunteers were heat treated for 30□min at 56□°C. Sixfold serially diluted plasma, from 1:3 to 1:2,430, were incubated with VACV-MVA, CPXV, or MPXV for 1□h at 37□°C. The mixture was subsequently incubated with HeLa (VACV-MVA) or BS-C-1 (CPXV & MPXV) in a 12-well plate for 1□h, for adsorption. Then, cells were overlayed with MEM supplemented with NaHCO_3_, 4%□FBS and 0.6% Avicel mixture. Plaques were resolved at 90□h post-infection by fixing in 10% formaldehyde for 1□h followed by staining in 0.5% crystal violet. All experiments were performed in parallel with sera from baseline controls, in an established viral concentration to generate 60–120 plaques per well.

### Poxvirus antigen sequence and structure analysis

The poxvirus antigen sequences were aligned using Clustal Omega and were colored based on sequence identity. The Mpox antigen structures of A29L, A35R, E8L and M1R were generated by SWISS homology model which used the solved PDB structures of Vaccinia antigens as templates. The PDB structure templates include PDB 4ETQ (E8L), 4M1G (A35R), 4U6H (M1R) and 3VOP (A29L). The protein structures were visualized in PyMol to simultaneously display protein backbone and surface with an emphasis on the neutralizing antibody epitopes. The antibody-antigen complex structures of A33R (PDB: 4M1G) and M1R (4U6H) were used to map the antibody binding sites on corresponding poxvirus antigens.

### Statistical Analysis

All analyses of patient samples were conducted using GraphPad Prism 10, JMP 15 and R 4.3.1. Multiple group comparisons were analyzed by running parametric (ANOVA) statistical tests. Multiple comparisons were corrected using Tukey’s and Dunnett’s tests as indicated in the figure legends. Single group comparisons were analyzed using Man-Whitney test.

## Acknowledgements

We are very grateful to all study participants who kindly donated specimens for this study. We also would like to thank D. Mucida who provided insight and expertise that greatly assisted the data analysis and comments that significantly improved this manuscript. We extend our gratitude to Nathan Smith Clinic group for their invaluable contributions to establishing the study platform and their dedicated engagement with the study participants. Additionally, we appreciate Julia Barret’s insightful discussions and input on this project. VSM is supported by the CAPES-YALE fellowship. Brazilian cohorts set up, sample collection and processing was supported in part by grants from Fundação Carlos Chagas Filho de Amparo à Pesquisa do Estado do Rio de Janeiro/FAPERJ (reference number E−26/210.785/2021) and from UFRJ (CIP: 24864/2022).

## Author Contributions

Conceived the study: LP, IY, SBO, and CL. Brazilian cohorts set up, sample collection and processing: LC, GM, GS, TMC, GSL, LMH, AMV. US cohort set up and sample collection: LP, IY, AZ, LB, SBC, CL. US cohort sample processing: VM, YZ, LL. Performed virus sequencing: MB, CBFV, and NDG. Cell culture and virus expansion: YZ, LL. Experiments and data collection: JC, VSM, LP, YZ, and CL. Data analysis: JC, VSM, AMV and CL. Virus Sequency identity analysis: ZF, SC. T cells Peptide pool design: AG, AS. Writing – Original draft: JC, VSM, ZF, and CL. Supervised the project: SBO, and CL. All authors reviewed and approved the manuscript.

## Competing interests

Alessandro Sette is a consultant for Gritstone Bio, Flow Pharma, Moderna, AstraZeneca, Qiagen, Fortress, Gilead, Sanofi, Merck, RiverVest, MedaCorp, Turnstone, NA Vaccine Institute, Emervax, Gerson Lehrman Group and Guggenheim. Alba Grifoni is a consultant for Sanofi and Pfizer. LJI has filed for patent protection for various aspects of T cell epitope and vaccine design work. All other authors declare no competing interests.

## Data availability

All the background information of participants and data generated in this study are included in Source Data Figure1. The genome information and aligned consensus genomes for poxviruses used in this study are available on NCBI (GenBank Accession numbers of Copenhagen (Cop)-D8L, A27L, A33R, L1R, B5R antigens of Mpox 2022 USA strain, Mpox Zaire-96-I-16 strain and Cowpox as well as their identity with corresponding antigens of Vaccinia virus WR strain are listed below. The Mpox 2022 genome (GCA_025462475.1) as well as reference genomes of Mpox Zaire strain (GCF_000857045.1), Vaccinia virus WR strain (GCF_000860085.1) and Cowpox (GCF_000839185.1) were used in the genome sequence alignment and analysis. Additional correspondence and requests for materials should be addressed to the corresponding author.

1. Vaccinia virus WR strain as sequence alignment references:

a. Cop-D8L: QTC35383.1, 100%
b. Cop-A27L: QTC35423.1, 100%
c. Cop-A33R: QTC35435.1, 100%
d. Cop-L1R: QTC35412.1, 100%
e. Cop-B5R: QTC35539.1, 100%
2. Mpox 2022 USA strain:

a. E8L, Cop-D8L: URK20542.1, 95%
b. A29L, Cop-A27L: URK20577.1, 94%
c. A35R, Cop-A33R: URK20584.1, 95%
d. M1R, Cop-L1R: URK20517.1, 99%
e. B6R, Cop-B5R: URK20605.1, 96%
3. Mpox Zaire-96-I-16 strain:

a. E8L, Cop-D8L: AAL40563.1, 94%
b. A29L, Cop-A27L: AAL40597.1, 94%
c. A35R, Cop-A33R: AAL40603.1, 95%
d. M1R, Cop-L1R: AAL40538.1, 99%
e. B6R, Cop-B5R: AAL40625.1, 96%
4. Cowpox:

a. CPXV125, Cop-D8L: ADZ30306.1, 98%
b. CPXV162, Cop-A27L: AAP48882.1, 98%
c. CPXV168, Cop-A33R: ADZ29703.1, 99%
d. CPXV99, Cop-L1R: ADZ24097.1, 100%
e. CPXV199, Cop-B5R: ADZ29304.1, 97%

## Extended Data Figures

**Extended Data Table 1.**
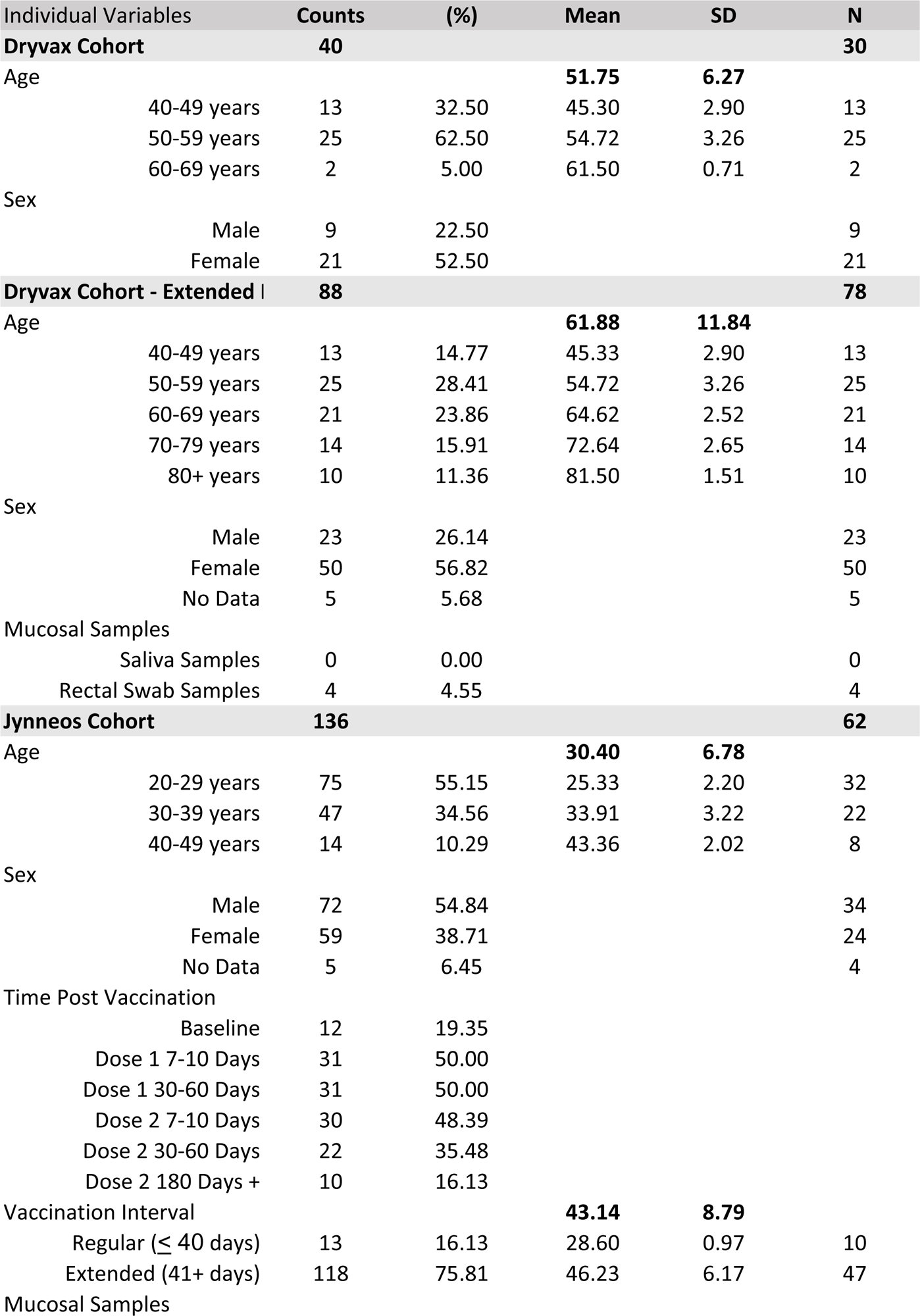
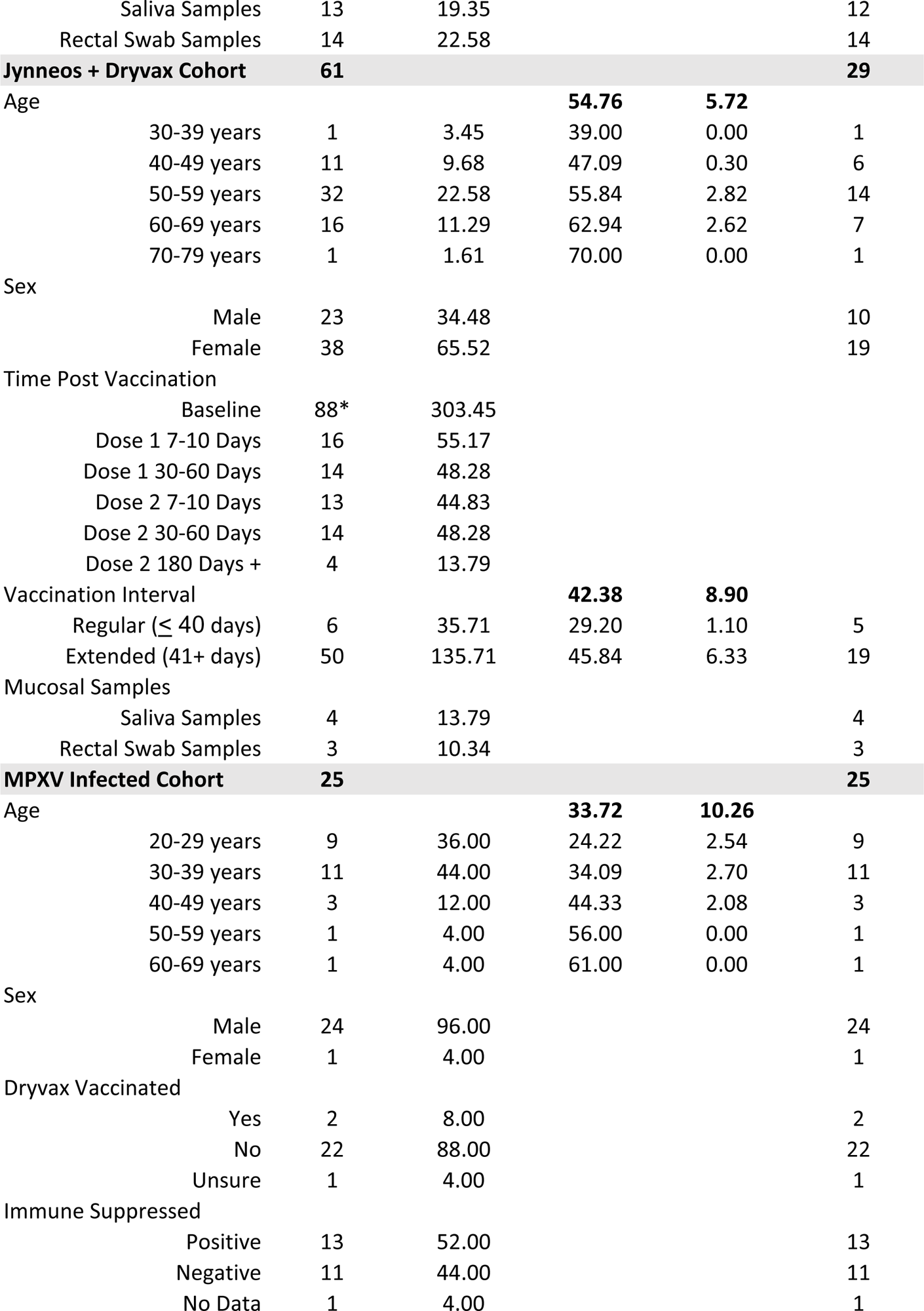
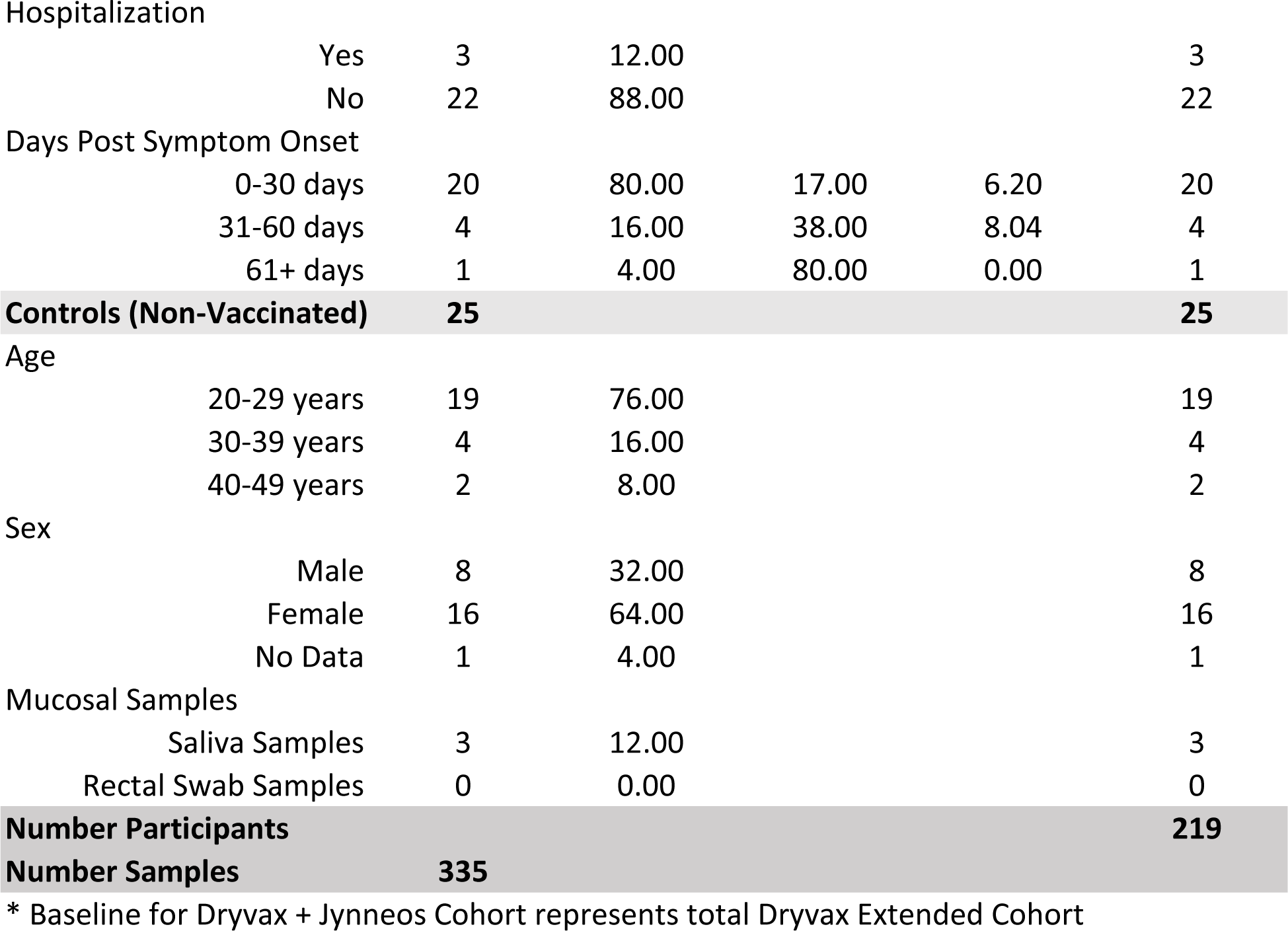
Detailed Clinical and Demographic Data for each Study Cohort. Demographic and relevant clinical data used within this study.

| Individual Variables |  | Counts | (%) | Mean | SD | N |
| --- | --- | --- | --- | --- | --- | --- |
| **Dryvax Cohort** |  | **40** |  |  |  | **30** |
| Age |  |  |  | **51.75** | **6.27** |  |
|  | 40-49 years | 13 | 32.50 | 45.30 | 2.90 | 13 |
|  | 50-59 years | 25 | 62.50 | 54.72 | 3.26 | 25 |
|  | 60-69 years | 2 | 5.00 | 61.50 | 0.71 | 2 |
| Sex |  |  |  |  |  |  |
|  | Male | 9 | 22.50 |  |  | 9 |
|  | Female | 21 | 52.50 |  |  | 21 |
| **Dryvax Cohort - Extended I** |  | **88** |  |  |  | **78** |
| Age |  |  |  | **61.88** | **11.84** |  |
|  | 40-49 years | 13 | 14.77 | 45.33 | 2.90 | 13 |
|  | 50-59 years | 25 | 28.41 | 54.72 | 3.26 | 25 |
|  | 60-69 years | 21 | 23.86 | 64.62 | 2.52 | 21 |
|  | 70-79 years | 14 | 15.91 | 72.64 | 2.65 | 14 |
|  | 80+ years | 10 | 11.36 | 81.50 | 1.51 | 10 |
| Sex |  |  |  |  |  |  |
|  | Male | 23 | 26.14 |  |  | 23 |
|  | Female | 50 | 56.82 |  |  | 50 |
|  | No Data | 5 | 5.68 |  |  | 5 |
| Mucosal Samples |  |  |  |  |  |  |
|  | Saliva Samples | 0 | 0.00 |  |  | 0 |
|  | Rectal Swab Samples | 4 | 4.55 |  |  | 4 |
| **Jynneos Cohort** |  | **136** |  |  |  | **62** |
| Age |  |  |  | **30.40** | **6.78** |  |
|  | 20-29 years | 75 | 55.15 | 25.33 | 2.20 | 32 |
|  | 30-39 years | 47 | 34.56 | 33.91 | 3.22 | 22 |
|  | 40-49 years | 14 | 10.29 | 43.36 | 2.02 | 8 |
| Sex |  |  |  |  |  |  |
|  | Male | 72 | 54.84 |  |  | 34 |
|  | Female | 59 | 38.71 |  |  | 24 |
|  | No Data | 5 | 6.45 |  |  | 4 |
| Time Post Vaccination |  |  |  |  |  |  |
|  | Baseline | 12 | 19.35 |  |  |  |
|  | Dose 1 7-10 Days | 31 | 50.00 |  |  |  |
|  | Dose 1 30-60 Days | 31 | 50.00 |  |  |  |
|  | Dose 2 7-10 Days | 30 | 48.39 |  |  |  |
|  | Dose 2 30-60 Days | 22 | 35.48 |  |  |  |
|  | Dose 2 180 Days + | 10 | 16.13 |  |  |  |
| Vaccination Interval |  |  |  | **43.14** | **8.79** |  |
| | Regular ($\leq 40$ days) | 13 | 16.13 | 28.60 | 0.97 | 10 |
|  | Extended (41+ days) | 118 | 75.81 | 46.23 | 6.17 | 47 |
| Mucosal Samples |  |  |  |  |  |  |

|  |  |  |  |  |  |
| --- | --- | --- | --- | --- | --- |
| Saliva Samples | 13 | 19.35 |  |  | 12 |
| Rectal Swab Samples | 14 | 22.58 |  |  | 14 |
| **Jynneos + Dryvax Cohort** | **61** |  |  |  | **29** |
| Age |  |  | **54.76** | **5.72** |  |
| 30-39 years | 1 | 3.45 | 39.00 | 0.00 | 1 |
| 40-49 years | 11 | 9.68 | 47.09 | 0.30 | 6 |
| 50-59 years | 32 | 22.58 | 55.84 | 2.82 | 14 |
| 60-69 years | 16 | 11.29 | 62.94 | 2.62 | 7 |
| 70-79 years | 1 | 1.61 | 70.00 | 0.00 | 1 |
| Sex |  |  |  |  |  |
| Male | 23 | 34.48 |  |  | 10 |
| Female | 38 | 65.52 |  |  | 19 |
| Time Post Vaccination |  |  |  |  |  |
| Baseline | 88* | 303.45 |  |  |  |
| Dose 1 7-10 Days | 16 | 55.17 |  |  |  |
| Dose 1 30-60 Days | 14 | 48.28 |  |  |  |
| Dose 2 7-10 Days | 13 | 44.83 |  |  |  |
| Dose 2 30-60 Days | 14 | 48.28 |  |  |  |
| Dose 2 180 Days + | 4 | 13.79 |  |  |  |
| Vaccination Interval |  |  | **42.38** | **8.90** |  |
| Regular ($\leq 40$ days) | 6 | 35.71 | 29.20 | 1.10 | 5 |
| Extended (41+ days) | 50 | 135.71 | 45.84 | 6.33 | 19 |
| Mucosal Samples |  |  |  |  |  |
| Saliva Samples | 4 | 13.79 |  |  | 4 |
| Rectal Swab Samples | 3 | 10.34 |  |  | 3 |
| **MPXV Infected Cohort** | **25** |  |  |  | **25** |
| Age |  |  | **33.72** | **10.26** |  |
| 20-29 years | 9 | 36.00 | 24.22 | 2.54 | 9 |
| 30-39 years | 11 | 44.00 | 34.09 | 2.70 | 11 |
| 40-49 years | 3 | 12.00 | 44.33 | 2.08 | 3 |
| 50-59 years | 1 | 4.00 | 56.00 | 0.00 | 1 |
| 60-69 years | 1 | 4.00 | 61.00 | 0.00 | 1 |
| Sex |  |  |  |  |  |
| Male | 24 | 96.00 |  |  | 24 |
| Female | 1 | 4.00 |  |  | 1 |
| Dryvax Vaccinated |  |  |  |  |  |
| Yes | 2 | 8.00 |  |  | 2 |
| No | 22 | 88.00 |  |  | 22 |
| Unsure | 1 | 4.00 |  |  | 1 |
| Immune Suppressed |  |  |  |  |  |
| Positive | 13 | 52.00 |  |  | 13 |
| Negative | 11 | 44.00 |  |  | 11 |
| No Data | 1 | 4.00 |  |  | 1 |
| Hospitalization |  |  |  |  |  |  |
|  | Yes | 3 | 12.00 |  |  | 3 |
|  | No | 22 | 88.00 |  |  | 22 |
| Days Post Symptom Onset |  |  |  |  |  |  |
|  | 0-30 days | 20 | 80.00 | 17.00 | 6.20 | 20 |
|  | 31-60 days | 4 | 16.00 | 38.00 | 8.04 | 4 |
|  | 61+ days | 1 | 4.00 | 80.00 | 0.00 | 1 |
| **Controls (Non-Vaccinated)** |  | **25** |  |  |  | **25** |
| Age |  |  |  |  |  |  |
|  | 20-29 years | 19 | 76.00 |  |  | 19 |
|  | 30-39 years | 4 | 16.00 |  |  | 4 |
|  | 40-49 years | 2 | 8.00 |  |  | 2 |
| Sex |  |  |  |  |  |  |
|  | Male | 8 | 32.00 |  |  | 8 |
|  | Female | 16 | 64.00 |  |  | 16 |
|  | No Data | 1 | 4.00 |  |  | 1 |
| Mucosal Samples |  |  |  |  |  |  |
|  | Saliva Samples | 3 | 12.00 |  |  | 3 |
|  | Rectal Swab Samples | 0 | 0.00 |  |  | 0 |
| **Number Participants** |  |  |  |  |  | **219** |
| **Number Samples** |  | **335** |  |  |  |  |

\* Baseline for Dryvax + Jynneos Cohort represents total Dryvax Extended Cohort

**Extended Data Figure 1.**
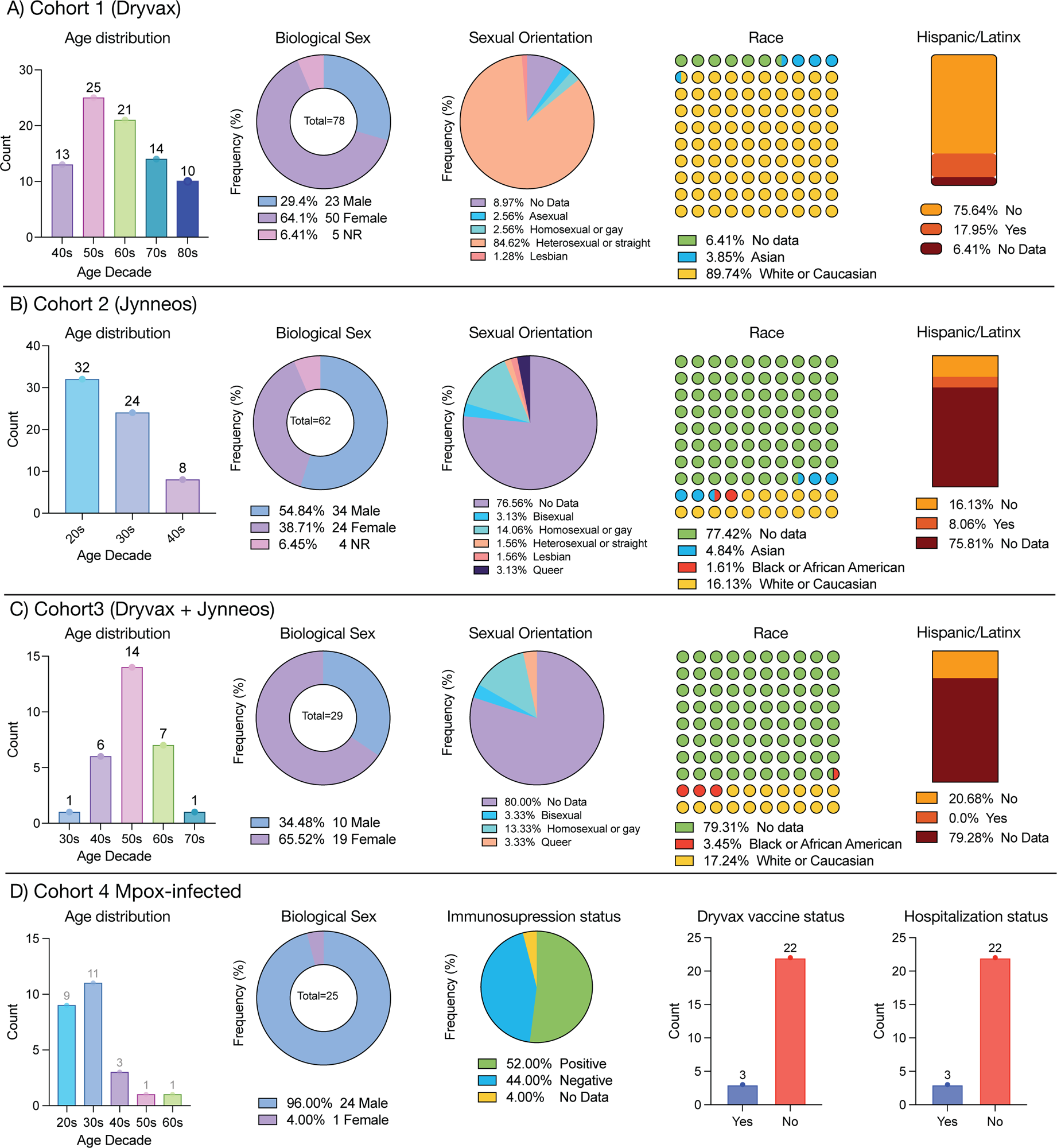
Demographics and relevant clinical data used within this study.

**Extended Data Figure 2.**
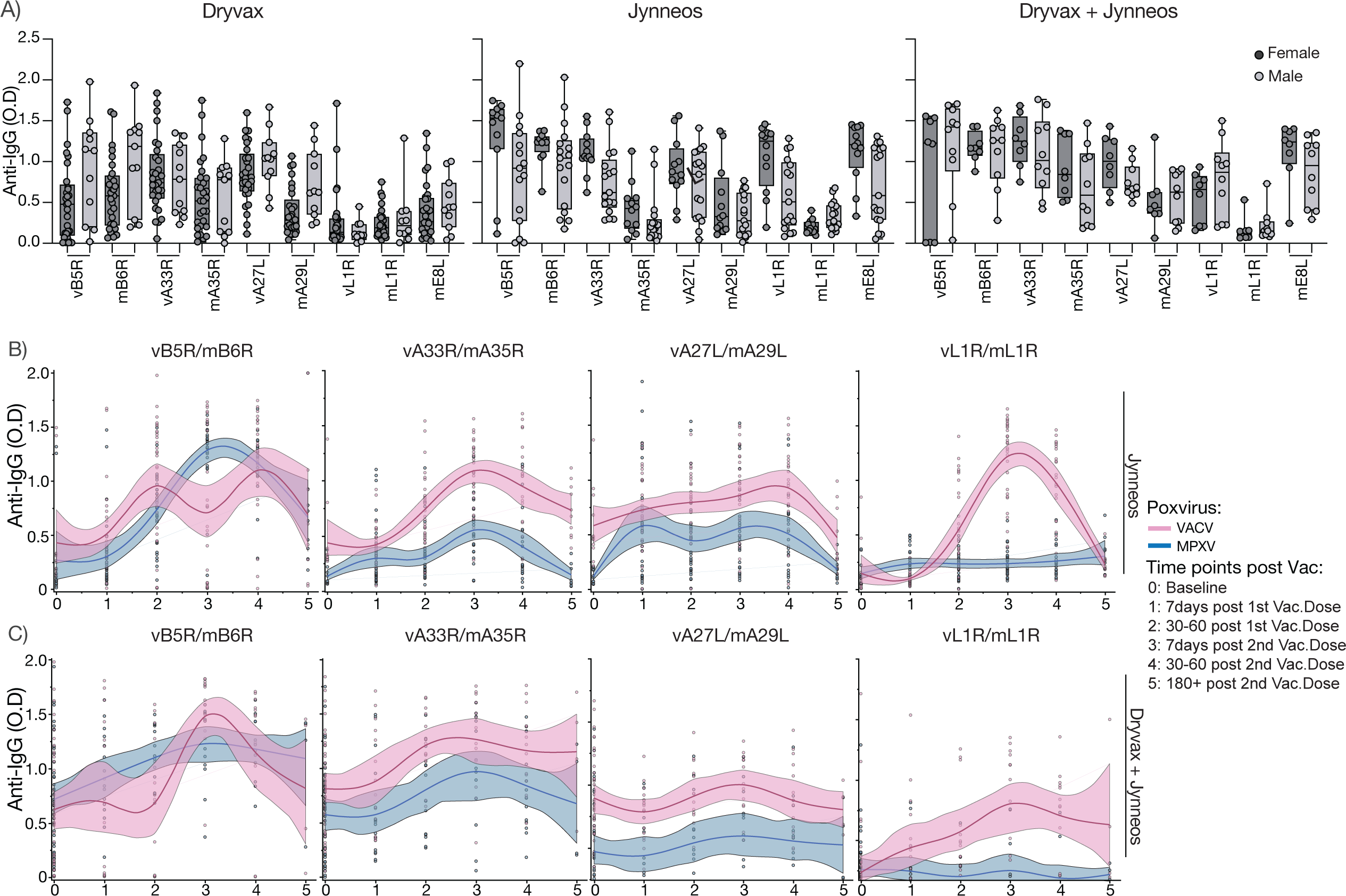
Comparison of plasma antibody reactivity to immunodominant primary antigens versus cross-reactive antigens. Vaccinated participants were stratified by poxvirus vaccination regimen and biological sex. Participants received Dryvax, JYNNEOS, or both Dryvax and JYNNEOS vaccines. Dryvax was administered as a single dose at least 40 years prior to this study, whereas the JYNNEOS regimen consisted of two vaccination doses and was recently administered to these participants. a, Plasma reactivity to VACV and MPXV proteins (B5/6R, A33/35R, A27/29L, L1/M1R, and E8L) in fully vaccinated individuals after stratification by biological sex. Female: Dryvax, n=29; JYNNEOS, n=12; Dryvax+JYNNEOS=8; Male: Dryvax, n=11; JYNNEOS, n=17; Dryvax+JYNNEOS=10. b,c, LOWESS regression analysis of virus-specific IgG levels over time following JYNNEOS vaccination. Regression lines indicate plasma IgG levels anti-VACV antigens (pink) or anti-MPXV antigens (blue); shading represents 95% confidence interval. For the JYNNEOS cohort, baseline controls consisted of non-vaccinated individuals. b, Virus-specific IgG levels over time in naïve, non-vaccinated individuals after the JYNNEOS regimen. c, Virus-specific IgG levels over time after the JYNNEOS regimen, in individuals previously vaccinated with Dryvax. For the JYNNEOS cohort, baseline controls consisted of non-vaccinated individuals. Baseline controls for the Dryvax + JYNNEOS cohort were individuals who had previously been vaccinated with Dryvax. Significance was assessed by one-way ANOVA corrected for multiple comparisons using Tukey’s method. Baseline: C2=12, C3=40; 7 days post 1^st^ vaccine dose: C2=31, C3=16; 30-60 days post 1^st^ vaccine dose C2=31, C3=14; 7 days post 2^nd^ vaccine dose: C2=30, C3=13; 30-60 days post 2^nd^ vaccine dose: C2=22, C3=14; 180-240 days post 2nd vaccine dose: C2=10, C3=4. Boxes represent average ± SD. TP5:180+, 180-240 days.

**Extended Data Figure 3.**
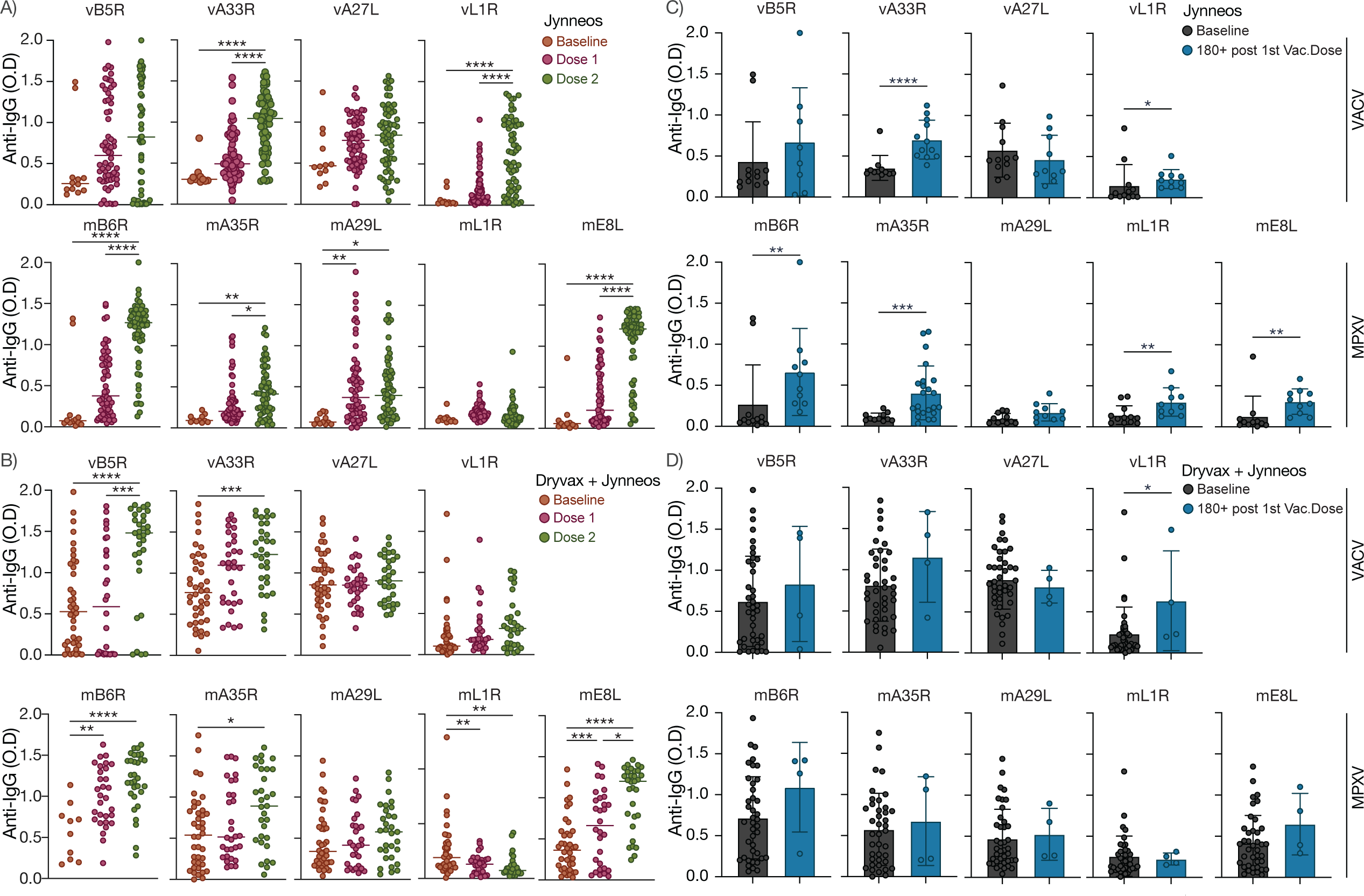
Dynamics of Plasma Antibody Titers Following JYNNEOS Vaccination doses. Participants received the JYNNEOS vaccine and plasma samples were collected longitudinally at multiple points: baseline (prior to the JYNNEOS vaccination), 7-30 days after the first vaccine dose, and 7-240 days after the second dose. The participants were then categorized into two groups based on their previous vaccination history: 1) the JYNNEOS group, consisting of naïve, non-vaccinated individuals who received the JYNNEOS regimen; and 2) the Dryvax + JYNNEOS group, comprising individuals who were previously vaccinated with Dryvax and subsequently received the JYNNEOS regimen. Importantly, for the JYNNEOS cohort, baseline consists of non-vaccinated individuals. Baseline controls for the Dryvax + JYNNEOS cohort were individuals who had previously been vaccinated with Dryvax. a,b,c,d Plasma reactivity to VACV proteins B5R, A33R, A27L, and L1 (top panel) and to MPXV proteins B6R, A35R, A29L, L1R and E8L (bottom panel). JYNNEOS: Baseline=12; Dose 1=62; Dose 2=62; JYNNEOS + Dryvax: Baseline=40; Dose 1= 30; Dose 2= 31. a, Plasma virus-specific IgG responses on naïve, non-vaccinated individuals following JYNNEOS vaccine doses. b, Plasma virus-specific IgG responses on individuals previously vaccinated with Dryvax and boosted with the JYNNEOS vaccine doses. c, Comparison of baseline plasma virus-specific IgG levels with those measured at the final collection time point (180-240 days post the second vaccination dose), in naïve, non-vaccinated individuals who received the JYNNEOS vaccine regimen. d, Comparison of baseline plasma virus-specific IgG levels with those measured at the final collection time point (180-240 days post the second vaccination dose) on individuals previously vaccinated with Dryvax and boosted with the JYNNEOS vaccine doses. Significance was assessed by one-way ANOVA corrected for multiple comparisons using Tukey’s method. JYNNEOS: Baseline=12; 180-240 days=10; JYNNEOS + Dryvax: Baseline=40; 180-240 days=4. Each dot represents a single individual. Horizontal bars represent median fold change. Boxes represent average ± SD. TP5:180+, 180-240 days. ****P < 0.0001, ***P < 0.001, **P < 0.01, and *P < 0.05.

**Extended Data Figure 4.**
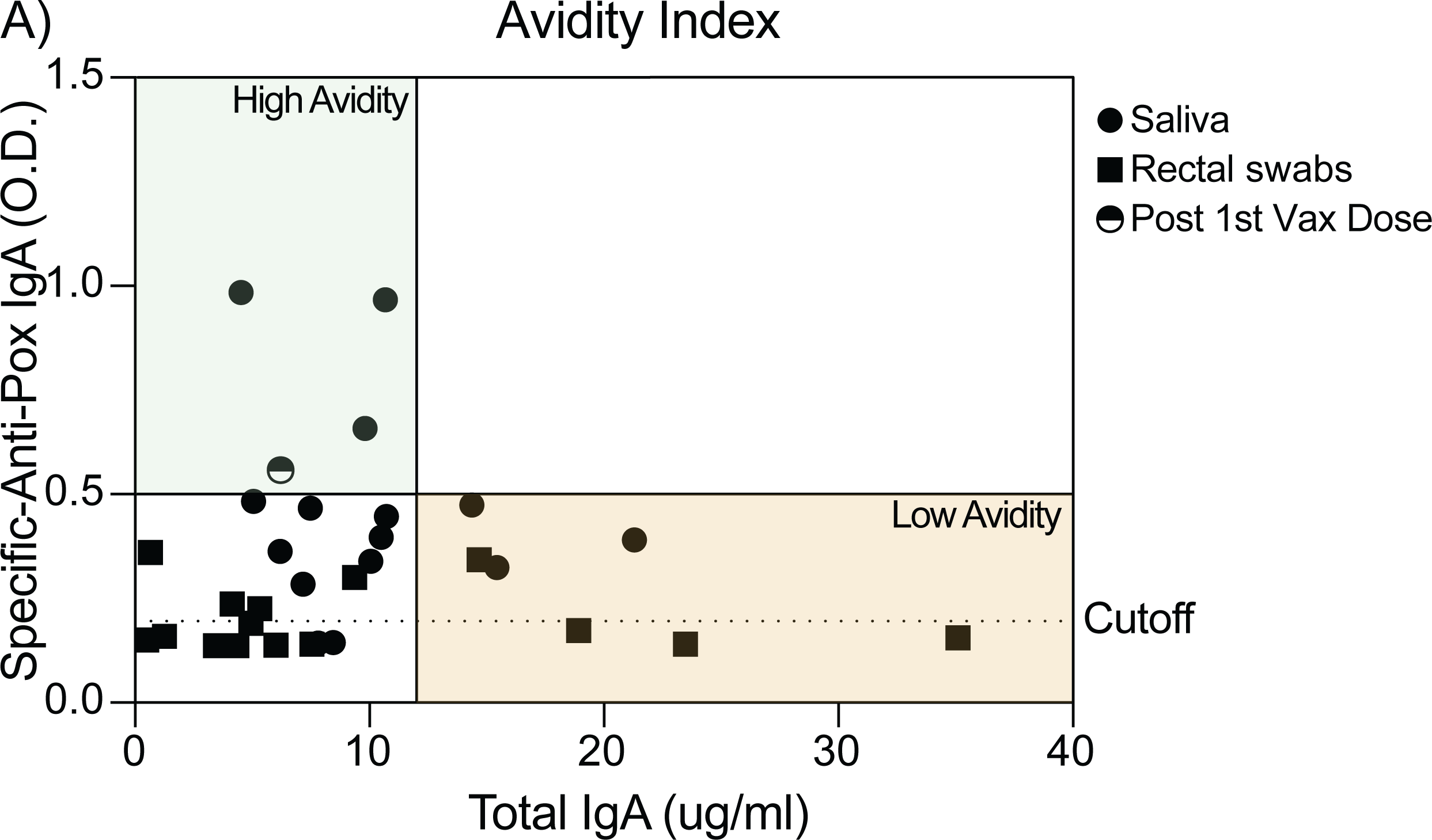
Avidity index for mucosal Orthopoxvirus-specific antibodies. Saliva and rectal swabs samples were collected following JYNNEOS vaccination and tested by ELISA for IgA levels against a mix of VACV and MPXV antigens (B5/6R, A33/35R, A27/29L, L1/M1R, and E8L). a, IgA avidity index was determined based on the ratio of specific anti-pox IgA to total IgA concentration. Green area denotes high antibody avidity area, while the yellow area indicates low avidity. Saliva samples were analyzed between 2 and 7 months post the second JYNNEOS vaccination dose, except for one sample, represented by a half-circle, which was analyzed 1 month post the first vaccination dose. Rectal swab samples were collected and analyzed 6-10 months following the second JYNNEOS dose. Saliva (n=17; C2=13; C3=4); rectal swab samples (n=21; C1=4; C2=14; C3=3). Cutoff was determined by calculating the mean of the non-vaccinated controls, n=3.

**Extended Data Figure 5.**
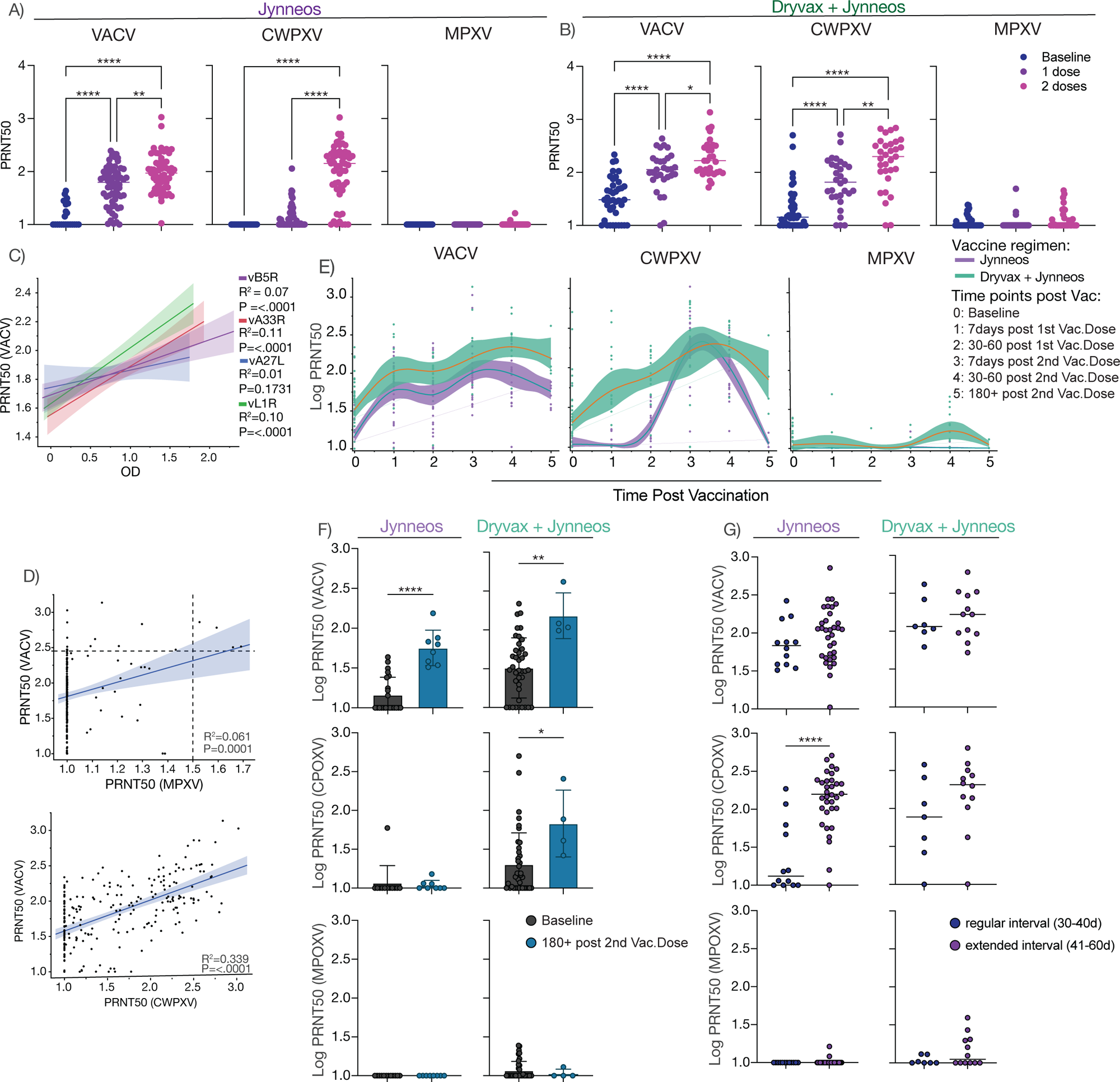
Dynamics of Plasma Neutralization Titers Following JYNNEOS Vaccination doses. Participants received the JYNNEOS vaccine and plasma samples were collected longitudinally at multiple points: baseline (prior to the JYNNEOS vaccination), 7-30 days after the first vaccine dose, and 7-240 days after the second dose. The participants were then categorized into two groups based on their previous vaccination history: 1) the JYNNEOS group, consisting of naïve, non-vaccinated individuals who received the JYNNEOS regimen; and 2) the Dryvax + JYNNEOS group, comprising individuals who were previously vaccinated with Dryvax and subsequently received the JYNNEOS regimen. Importantly, for the JYNNEOS cohort, baseline consists of non-vaccinated individuals. Baseline controls for the Dryvax + JYNNEOS cohort were individuals who had previously been vaccinated with Dryvax. Analysis of immunogenicity were performed using neutralization assays with authentic Vaccinia virus (VACV), Cowpox virus (CWPXV) and Mpox virus (MPXV) virus. a,b, Plasma neutralization titers against VACV, CWPXV and MPXV following JYNNEOS vaccination doses. **a,** Neutralization capacity in naïve, non-vaccinated individuals following JYNNEOS vaccine doses, Baseline, n=22; 1 Dose, n=62; 2 Doses=60. **b,** Neutralization capacity on individuals previously vaccinated with Dryvax and boosted with the JYNNEOS vaccine doses, Baseline, n=40; 1 Dose, n=30; 2 Doses=31. **c,** Linear regression analysis assessing the relationship between the neutralization titers against VACV and the IgG levels to VACV as determined by ELISA antigens. Regression lines are shown as blue purple (vB5R), red (vA33R), (vA27L) or green (vL1R), or Pearson’s correlation coefficients and linear regression significance are listed accordingly; shading represents 95% confidence interval. **d,** Linear regression analysis assessing the relationship between the neutralization titers against VACV and neutralization titers against MPXV (top) and CWPXV (bottom). Pearson’s correlation coefficients and linear regression significance are listed accordingly; shading represents 95% confidence interval. **e,** Plasma neutralization capacity against VACV, CWPXV and MPXV over time following JYNNEOS vaccination. **f,** Comparison of baseline neutralization titers with those measured at the final collection time point (180-240 days post the second vaccination dose), in both naïve, non-vaccinated individuals and those previously vaccinated with Dryvax who received the JYNNEOS vaccine regimen. JYNNEOS: Baseline=12; 180-240 days=8; Dryvax: Baseline=40; 180-240days=4. **g,** Neutralization capacity against VACV, CWPXV, and MPXV in participants that received the 2^nd^ JYNNEOS vaccine dose at the regular interval, 4-5 weeks (C2, n=12; C3, n=7) or an extended interval, 7-8 weeks (C2, n=34; C3, n=12). Significance was assessed by a non-parametric Mann-Whitney t test. Each dot represents a single individual, values chosen for this analysis were closest to peak of antibody response (7-30 days post second dose). Horizontal bars represent average ± SD. Boxes represent average ± SD. TP5:180+, 180-240 days. ****P < 0.0001, ***P < 0.001, **P < 0.01, and *P < 0.05.

**Extended Data Figure 6.**
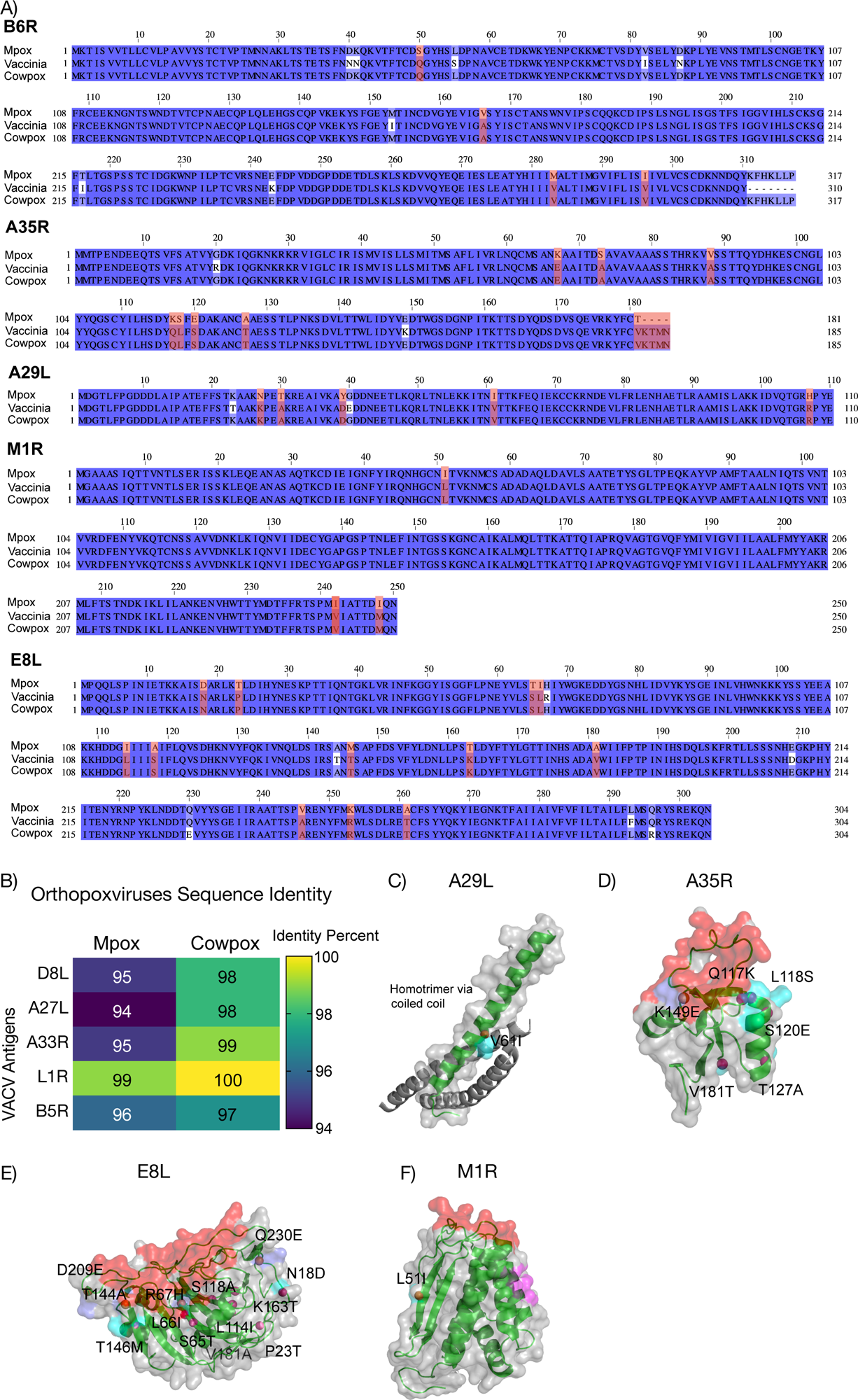
Sequence identity and structural analysis of Orthopoxviruses neutralizing antibody targets. **a,** Comparative analysis of conserved amino acid sequences among MPXV, VACV, and CWPXV within immunogenic proteins B6R, A35R, A29L, M1R, and E8L. Amino acids highlighted in white indicate point mutations unique to VACV antigens. Amino acids highlighted in red represent point mutations exclusive to MPXV. **b,** Heat map analysis depicting the percentage of sequence identity between MPXV and CWPXV compared to VACV’s primary neutralization targets proteins. **c,d,** Structural models of Mpox antigens, based on Vaccinia antigen structures. Neutralizing antibody binding sites are highlighted in red. MPXV specific residues are highlighted in blue, and MPXV/CPXV specific residues are highlighted in purple. Unique mutations specific to MPXV are indicated by the red dots.

**Extended Data Figure 7.**
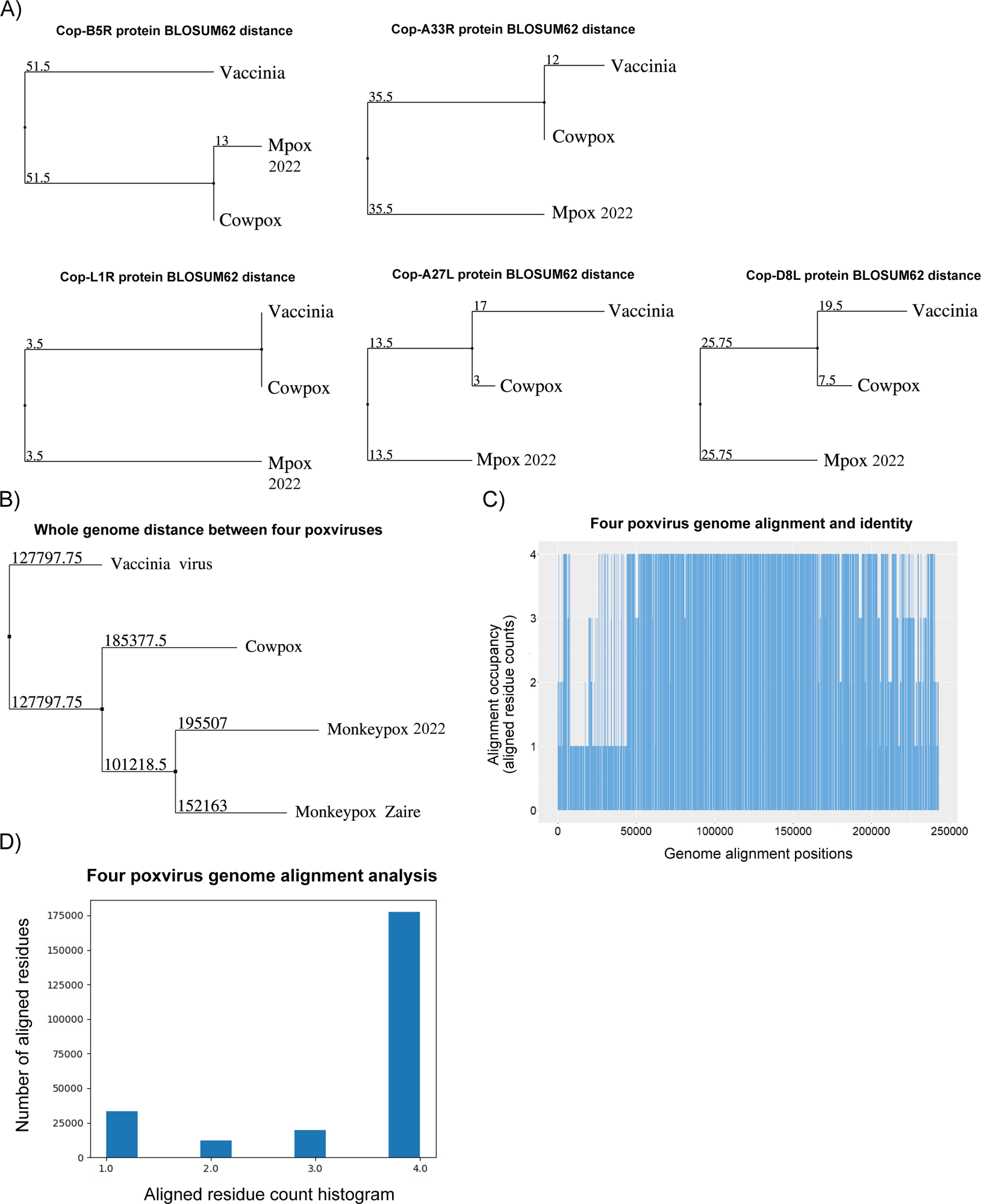
Similarity analysis of key antigen protein sequence and whole genome DNA sequence between Vaccinia virus, Cowpox, Mpox 2022 and Mpox Zaire strain. **a,** Among the tripartite distance relationships, conserved poxvirus antigens showed closest protein distance between Vaccinia virus and Cowpox, except for B5R that exhibited closest distance between Cowpox and Mpox. The protein distance was calculated using the BLOSUM62 matrix and the phylogenetic tree was plotted using neighbor joining method. **b,** The whole genomes of four poxviruses were aligned and their DNA distance or conservation was calculated and shown in the phylogenomic tree. Cowpox genome showed a higher sequence identity with Mpox 2022 or Mpox Zaire strain as compared to vaccinia virus genome. **c,** The identical nucleotide counts were plotted on the aligned positions of four poxvirus genomes (minimum 1, maximum 4). d, Histogram showing the number of aligned positions in each categories of identical nucleotide counts based on the genome alignment in Extended Data Fig. 7c.

## Notes

### Funding Statement

SBO and CL secured funding for this study. VSM is supported by the CAPES-YALE Fellowship. Brazilian cohorts set up, sample collection and processing was supported in part by grants from Fundacao Carlos Chagas Filho de Amparo a Pesquisa do Estado do Rio de Janeiro/FAPERJ and from UFRJ.

### Author Declarations

The Human Research Protection Program Institutional Review Board of Yale University gave ethical approval for this work (IRB protocol ID 2000033415). The Research Ethics Committee of the Hospital Universitario Clementino Fraga Filho/Universidade Federal Rio de Janeiro gave ethical approval for this work (protocol number CAAE 62281722.5.0000.5257).

